# Data Extraction from Oncology Imaging Reports by Large Language Models: A Comparative Accuracy Study

**DOI:** 10.64898/2025.12.30.25343206

**Authors:** Lea P. Passweg, Johannes M. Schwenke, Christof M. Schönenberger, Flavio Locher, Julia Picker, Manuel Dieterle, Benjamin Thiele, Dimitri Hasler, Alessia Danelli, Andreas M. Schmitt, Tobias Heye, Thomas Stojanov, Matthias Briel, Benjamin Kasenda

## Abstract

**Importance:** Manual data extraction from clinical text is resource intensive. Locally hosted large language models (LLMs) may offer a privacy-preserving solution, but their performance on non-English data remains unclear.

**Objective:** To investigate whether the classification accuracy of locally hosted LLMs is non-inferior to human accuracy when determining metastasis status and treatment response from German radiology reports.

**Design:** In this retrospective comparative accuracy study, five locally hosted LLMs (llama3.3:70b, mistral-small:24b, qwq:32b, qwen3:32b, and gpt-oss:120b) were compared against humans. To calculate accuracy, a ground truth was established via duplicate human extraction and adjudication of discrepancies by a senior oncologist. Both initial human extraction and LLM outputs were compared against this ground truth.

**Setting:** The study was conducted at a tertiary referral hospital in Switzerland; data processing and analyses took place inside the hospital network.

**Participants:** 400 randomly sampled radiology reports from adult cancer patients (CT, MRI, PET) generated between January 2023 and May 2025.

**Exposures:** Automated classification of metastasis status and treatment response by LLMs using a standardized prompt pipeline compared to manual human review.

**Main Outcomes and Measures:** Primary outcomes were non-inferiority (5 percentage points [pp] margin) of LLM classification accuracy compared with human accuracy for metastasis status (presence/absence by anatomical site) and treatment response categories. Secondary outcomes included accuracy for primary tumor diagnosis, radiological absence of tumor, and extraction time per report.

**Results:** The analysis included 400 reports from 317 patients (mean age 63 years, 32% women). On the test set (n=300), human accuracy for metastasis status was 98.4% (95% CI 98.0%–98.8%). All LLMs were non-inferior; gpt-oss:120b performed best (97.6% accuracy; difference:xs −0.8pp [90% CI, −1.3 to −0.3 pp]). For response to treatment, human accuracy was 86.0% (95% CI 83.2%–88.8%). All LLMs were inferior; the most accurate model, gpt-oss:120b, achieved 78.3% (difference −7.7 pp [90% CI, −11.6 to −3.8 pp]). Mean human time per report was 120 seconds vs 11–63 seconds for LLMs.

**Conclusion and Relevance:** In this study, LLMs were non-inferior to human accuracy for classification of metastasis status but were inferior for response to treatment assessment. gpt-oss:120b was the most accurate among tested LLMs.

**Study Registration:** OSF: 45PVQ

**Key Points:** *Question:* Can locally hosted large language models (LLMs) match human performance when extracting sites of metastases and response to treatment from radiology reports of cancer patients?

*Findings:* In this preregistered, single center study of 300 German radiology reports, all evaluated LLMs were non-inferior to humans in extracting the presence or absence of metastasis by organ site, but LLMs were inferior to humans in classification of response to treatment.

*Meaning:* LLMs can be suitable for classification of metastasis status, whereas more caution is warranted for more complex tasks where additional clinical reasoning may be required.

## Introduction

Structured clinical data are essential for quantitative research, including descriptive statistics, prediction modeling, and clinical trials. Valuable clinical information, such as information on treatment response in cancer patients, is typically embedded in unstructured free-text clinical notes in electronic health records, such as radiology or consultation reports. Manual extraction of these data is resource-intensive, error-prone, and depends on training of data coders ^1,2^. This challenge is particularly critical in oncological studies, where radiological treatment response evaluation frequently serves as a primary outcome.

Recent advances in large language models (LLMs) have shown potential in data extraction and classification ^3–5^. Locally hosted LLMs preserve privacy and security compared to closed-sourced alternatives, as sensitive patient data remain within a health service provider’s secure IT infrastructure ^6^. It is unclear how well locally hosted LLMs perform on real-world, particularly non-English, radiology reports of cancer patients and whether their outputs are reliable enough to support research and clinical workflows.

We tested a secure, LLM-based, automated pipeline which extracts information on primary tumor diagnosis, metastasis location, and response to treatment of cancer patients from radiology reports. To assess the accuracy of our automated pipeline, we compared data extraction by humans with data extracted using our pipeline.

## Methods

### Study design and setting

This retrospective study was conducted at the University Hospital Basel, Switzerland. All data processing occurred within the secure hospital network. Extracted data were stored in a locally hosted REDCap database ^7^.

### Radiology reports

German radiology reports were retrieved from the Clinical Data Warehouse (CDWH) of the hospital; the data flow is shown in eFigure 1. We included reports from computed tomography (CT), magnetic resonance imaging (MRI), and positron emission tomography-CT (PET-CT) ordered by physicians of the Division of Medical Oncology between January 2023 and May 2025, in adult (≥18 years) patients with cancer who had provided the hospital’s General Research Consent ^8^. To allow meaningful response assessment and minimize reports on baseline-staging, exams had to occur ≥2 months after the first outpatient oncology appointment. Reports clearly irrelevant for tumor-response assessment according to their CDWH identifier, such as reports on imaging-guided interventions, were excluded. Based on a power simulation, we randomly sampled 400 imaging reports and split the sample into a prompt-optimization set (n=100) and test set (n=300).

### Extraction tasks

The data extraction comprised four tasks: (1) classification of primary tumor diagnosis form the medical history embedded in the radiology report as one of 28 prespecified SNOMED Clinical Terms categories (e.g. colorectal cancer); (2) classification of metastasis status, as present or absent for each metastasis region (e.g., lung, liver, bone, brain, lymph nodes), depending on whether the region of interest was relevant for the respective imaging exam; (3) classification of treatment response (on PET-CT as complete response, partial response, stable disease, progressive disease, or not applicable; on CT/MRI as response, stable disease, progressive disease, not applicable). The ‘not applicable’ category was assigned when the imaging study was not performed for treatment monitoring (e.g., baseline staging). (4) classification of radiological absence of any tumor (yes/no). We *a priori* decided against an attempt to differentiate between complete response and partial response of reports from CT or MRI, based on clinical experience that this distinction can be challenging. We did not categorize responses according to the Response Evaluation Criteria in Solid Tumors (RECIST) or the Response Assessment in Neuro-Oncology (RANO), because these are not routinely applied and reported outside of clinical trials.

### Ground truth

Two groups of human coders (medical master students and medical doctors) independently extracted data in duplicate, using the same written guidelines (see eMethods). Disagreements were adjudicated by a senior oncologist, establishing the ground truth. Inter-rater agreement between the two human coders was assessed using raw percent agreement and Cohen’s Kappa (κ).^9^ To assess the robustness of this human-derived ground truth, we performed a *post hoc* sensitivity analysis on the test set. We identified all instances where the best-performing model disagreed with the ground truth for the primary outcomes. These discrepancies were reviewed by two senior oncologists (BK, AMS) who were blinded to the source of the extraction. This review established a “revised ground truth” used specifically for the sensitivity analysis.

### LLMs and prompting

We evaluated five locally hosted LLMs using the hospital’s graphics processing unit (GPU) cluster. The cluster consists of a high-density GPU server equipped with dual 48-core x86 processors, approximately 2.3 TB of DDR5 system memory, and eight NVIDIA H200 PCIe Gen5 GPUs, each providing 141 GB of high-bandwidth memory. Models were locally deployed using ollama version 12.2 and accessed through R using ellmer version 0.3.0 ^10,11^. We evaluated the performance of the following models: llama3.3:70b (full name: llama3.3:70b-Instruct-q5_K_M, knowledge cutoff December 2023, release date December 6, 2024) ^12^, mistral-small:24b (full name: mistral-small:24b-Instruct-2501-q4_K_M, knowledge cutoff October 2023, release date January 30, 2025) ^13^, qwq:32b (knowledge cutoff November 2024, release date March 5, 2025), qwen3:32b (knowledge cutoff not stated, release date April 29, 2025) ^14^ and gpt-oss:120b (knowledge cutoff September 2024, release date August 5, 2025) ^15^. Models were used without additional fine-tuning and prompted in English.

Prompts were iteratively refined on the prompt-optimization set (n=100) and applied to the test set (n=300). We implemented a two-step pipeline where models provided free-text reasoning for generating structured JSON. This led to improvements in performance for the extraction of tasks across all models except gpt-oss:120b which required direct output (see eFigure 2, eMethods). Prompts were adapted to the imaging exam (PET-CT vs CT/MRI; body region of the imaging exam; see eFigures 3–4). Inference settings were fixed across models with a context window of 10,000 tokens and a temperature of 0 to maximize reproducibility. For the exact prompts and implementation details, see eMethods and code on GitHub ^16^.

### Outcomes

The two prespecified primary outcomes were the difference in human versus LLM accuracy for classification of metastasis status and treatment response assessment ^17^. Secondary outcomes were the accuracy for classification of the primary diagnosis, radiological absence of tumor, and overall accuracy across all four tasks, as well as the mean time of extraction per report. Exploratory analyses included sensitivity, specificity, negative and positive predictive values.

### Statistical analysis

For each outcome, the initial human extractions and responses from each LLM were compared against the established ground truth. To compare the performance of LLMs with humans, we used logistic regression models to estimate quantities of interest. We accounted for within-report correlation using cluster-robust standard errors at the report level ^18,19^. We tested for non-inferiority in the primary outcomes against a pre-specified margin of −0.05 for the difference in accuracy. This margin served as a pragmatic feasibility threshold to determine if model performance warranted further development, rather than a strict clinical safety limit. Standard errors were estimated using the delta method ^20(chap14)^. For exploratory outcomes (sensitivity, specificity, negative predictive value [NPV], positive predictive value [PPV]), we used bootstrapping, with 1000 repetitions, as the delta method estimates produced confidence intervals crossing 0 or 1. We did not adjust for multiplicity across LLMs and tasks. To check the robustness of our findings, we performed a Bayesian sensitivity analysis with weakly informative priors.

We estimated the human time-per-report using REDCap audit logs, which timestamped when a data extraction form was saved. For each user, we calculated the time gap (in seconds) between consecutive saves. Gaps exceeding 300 seconds were classified as breaks and were treated as missing values. These missing values were then imputed using the mean of all observed gaps across the dataset. Mean gap time served as the approximate human time-per-report. For the LLMs, we recorded the total processing time for all reports and calculated the mean time-per-report. All analyses were performed in R (version 4.4.3) using marginaleffects (version 0.30.0) for model-based predictions, and the tidyverse (version 2.0.0) for data handling and visualization ^21–23^.

### Sample size rationale

We conducted a power simulation ^16^. For metastasis classification, with a conservative estimate of 3 metastasis locations to classify as absent or present per report, we estimated our power is to be >95% to detect non-inferiority at an α of 0.05, if the true LLM accuracy were to equal human accuracy. For classification of treatment response assessment, our estimated power is 70–90%, depending on report heterogeneity. Overall, 300 reports provide high power for metastasis classification and adequate power for the response to treatment.

## Results

### Sample Characteristics

We included a total of 400 radiology reports from 317 patients (mean age 63 ± 15 years; 32% women). Most reports were CT and MRI (89%, n=355), with the remaining 11% (n=45) being PET-CT scans (Table 1).

**Table 1:** Patient characteristics, imaging modality, primary diagnosis, metastasis location, treatment response on CT/MRI and PET–CT, and tumor-free status of the study cohort (n=400). Abbreviations: PET–CT, positron emission tomography–computed tomography; CT, computed tomography; MRI, magnetic resonance imaging; y, years; SD, standard deviation. *Other primary tumors comprise lymphoma: 33 (8%), prostate cancer: 26 (7%), sarcoma: 23 (6%), head and neck tumors: 18 (5%), cancer of the urinary tract (including ureter and bladder): 13 (3%), other: 8 (2%), testicular cancer: 8 (2%), no malignant disease: 6 (2%), kidney tumor: 5 (1%), unclear: 2 (1%), breast cancer: 1 (<1%), cancer of unknown primary: 1 (<1%), thyroid cancer: 1 (<1%). **Other metastasis locations comprise soft tissue: 26 (5%), cns: 20 (4%), pleura: 18 (4%), peritoneum: 14 (3%), adrenal: 12 (2%), pancreas: 8 (2%), spleen: 6 (1%), kidney: 4 (1%).

| Patient Characteristics |  |
| --- | --- |
| Age, mean (SD), y | 63.28 (15.1) |
| Female Sex | 103 (32%) |
| Imaging modality |  |
| PET–CT | 45 (11%) |
| CT/MRI | 355 (89%) |
| Primary Diagnosis |  |
| Lung Tumor (including Mesothelioma) | 95 (24%) |
| Gastrointestinal Tumors | 63 (16%) |
| Primary Brain Tumor | 56 (14%) |
| Primary Skin Tumor (including Melanoma and Non-Melanoma) | 41 (10%) |
| Other* | 145 (36%) |
| Metastasis location |  |
| Lymph nodes | 132 (33%) |
| Bone | 108 (27%) |
| Lung | 96 (24%) |
| Liver | 68 (17%) |
| Other** | 108 (27%) |
| Treatment response (CT/MRI) (n=355) |  |
| Response | 174 (49%) |
| Stable Disease | 80 (23%) |
| Progression | 73 (21%) |
| Not applicable | 28 (8%) |
| Treatment response (PET–CT) (n=45) |  |
| Complete Response | 19 (42%) |
| Partial Response | 11 (24%) |
| Progression | 11 (24%) |
| Not applicable | 3 (7%) |
| Stable Disease | 1 (2%) |
| Tumor-free status |  |
| No | 244 (61%) |
| Yes | 125 (31%) |
| Not applicable | 31 (8%) |

### Human inter-rater Agreement

Across the 400 radiology reports, the two human coders independently classified a total of 4,669 data items in duplicate. This comprised 3,469 assessments of specific metastasis sites and 400 classifications each for primary diagnosis, treatment response, and radiological tumor-free status (no evidence of disease). Agreement was 91.5% for primary diagnosis (κ = 0.90), 96.6% for metastasis status (κ = 0.76), 80.5% for radiological tumor-free status (κ = 0.63), and 76.2% for response to treatment (κ = 0.68).

### Metastasis Status

Performance metrics were calculated exclusively on the test set (n=300 reports). The ground truth for metastasis classification (presence vs absence by site) contained 193 positive and 2441 negative items. Human accuracy compared to ground truth was 98.4% (95% CI 98.0%–98.8%). All LLMs met the prespecified non-inferiority margin. gpt-oss:120b had the highest accuracy among LLMs (97.6%; 95% CI 96.9%–98.2%), followed closely by the other models (range: 95.7%–97.4%) (Table 2, Figure 1, eFigure 5).

**Table 2:** Table shows pooled accuracy and 95% confidence intervals (CIs) for humans and LLMs, as well as the difference in accuracy (LLM accuracy − human accuracy) with 90% CIs and corresponding one-sided non-inferiority p-values. The upper panel reports binary classification of site-specific metastases (present vs. absent). The lower panel reports multi-class classification of response to treatment.

| Model | Accuracy | Difference from Human | p-value |
| --- | --- | --- | --- |
|  | % (95% CI) | Percentage Points (90% CI) | Non-inferiority |
| Metastasis |  |  |  |
| human | 98.4% (98.0%–98.8%) | [Reference] |  |
| gpt-oss:120b | 97.6% (96.9%–98.2%) | -0.8 (-1.3 to -0.3) | <0.001 |
| qwen3:32b | 97.0% (96.3%–97.6%) | -1.4 (-2.0 to -0.8) | <0.001 |
| qwq:32b | 97.4% (96.7%–98.0%) | -1.0 (-1.5 to -0.5) | <0.001 |
| mistral-small:24b | 95.8% (95.0%–96.6%) | -2.6 (-3.2 to -1.9) | <0.001 |
| llama3.3:70b | 95.7% (94.9%–96.6%) | -2.6 (-3.3 to -1.9) | <0.001 |
| Response to Treatment |  |  |  |
| human | 86.0% (83.2%–88.8%) | [Reference] |  |
| gpt-oss:120b | 78.3% (73.7%–83.0%) | -7.7 (-11.6 to -3.8) | 0.869 |
| qwen3:32b | 72.0% (66.9%–77.1%) | -14.0 (-18.1 to -9.9) | >0.999 |
| qwq:32b | 73.7% (68.7%–78.7%) | -12.3 (-16.3 to -8.4) | 0.999 |
| mistral-small:24b | 65.9% (60.5%–71.3%) | -20.1 (-24.5 to -15.8) | >0.999 |
| llama3.3:70b | 69.3% (64.1%–74.6%) | -16.7 (-20.9 to -12.4) | >0.999 |

**Figure 1:**
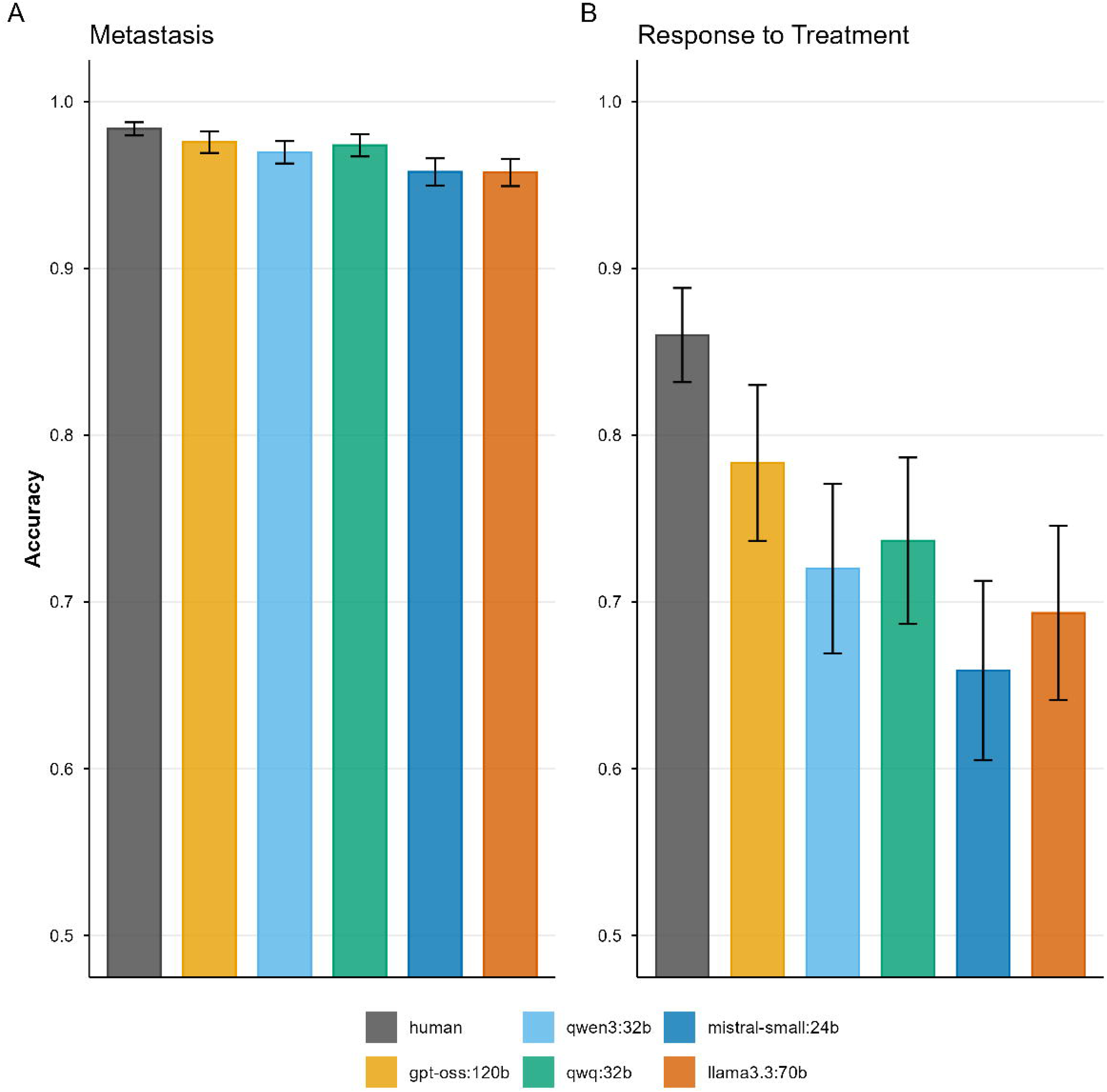
Plots show pooled accuracy and 95% confidence intervals (CIs) of humans and LLMs. **(A)** Binary classification of site-specific metastases (present vs. absent). **(B)** Multi-class classification of response to treatment. (Categories for CT/MRI: stable disease, response, remission, not applicable; for PET: stable disease, partial response, complete response, remission, not applicable).

In exploratory analyses, human coders achieved a PPV of 86.9% (95% CI 83.1%–90.8%) and NPV of 99.3% (95% CI 99.1%–99.6%). LLM performance varied: gpt-oss:120b was conservative with a high PPV of 90.6% (95% CI 86.0%–95.1%) and lower NPV (98.0%; 95% CI 97.4%–98.7%). In contrast, llama3.3:70b had a low PPV (64.6%; 95% CI 58.3%–70.9%) with an NPV of 99.4% (95% CI 99.1%–99.7%) (eFigures 6–7, eTable 1). Errors clustered in lung and lymph nodes (Figure 2), where gpt-oss:120b produced predominantly false negatives, whereas llama3.3:70b produced predominantly false positives.

**Figure 2:**
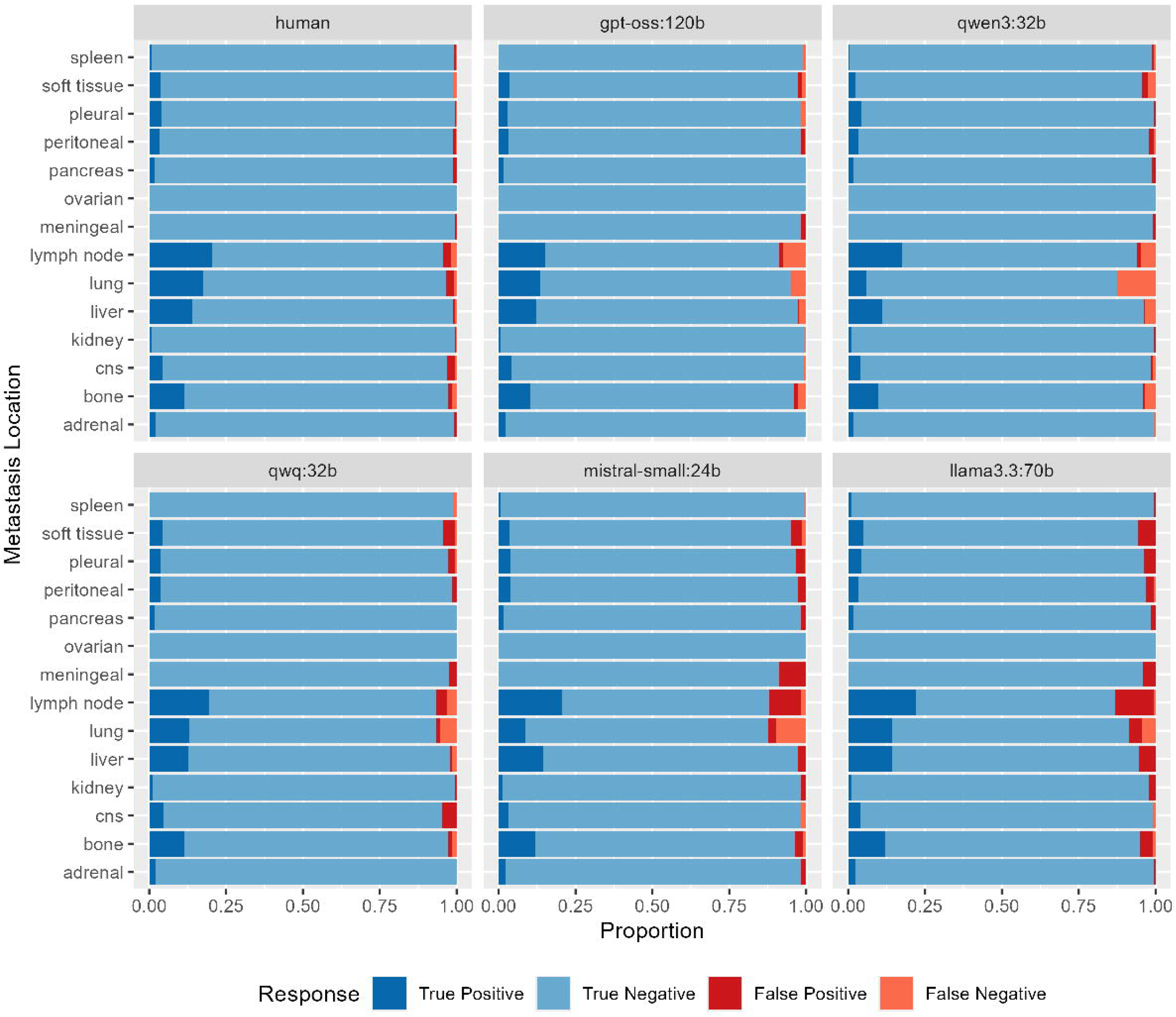
The plot shows the distribution of classification errors of metastasis status across anatomical sites, stratified by human coders and LLMs. Classification of findings in the lung and lymph nodes were the primary errors of humans. This was expected, given the more ambiguous language frequently used to describe findings in these sites. Interestingly, LLMs mainly struggled in the same areas. LLMs show different diagnostic profiles: for example, gpt-oss:120b and qwen3:32b appear to classify conservatively (higher threshold to classify a finding as metastasis), whereas llama3.3:70b and mistral-small:24b appears to classify findings as metastasis more liberally. Abbreviations: CNS, central nervous system.

### Response to Treatment

For assessment of treatment response, human accuracy was 86.0% (95% CI 83.2%–88.8%). No LLM achieved non-inferiority. The best-performing model, gpt-oss:120b, achieved 78.3% accuracy (95% CI 73.7%–83.0%) (Table 2, Figure 1, eFigure 5).

When assessing treatment response by classification category in exploratory analyses, accuracy varied substantially (eFigure 8–9). LLMs underperformed humans within the *progression* or *stable disease* categories. For *progression* humans were 96.7% accurate (95% CI 93.1%–99.3%); the best performing models, mistral-small:24b and qwq:32b, achieved 77.4% (95% CI 67.6%–86.2%), with other models ranging between 70.0% and 73.4%. For *stable disease* human accuracy was 74.1% (95% CI 65.5%–82.1%); the best performing model, gpt-oss:120b, achieved 58.6% (95% CI 46.5%–70.0%), while other models fell below 50% (eTable 2).

To assess the clinical utility, we simplified the task to a binary classification (progression vs. non-progression). In this setting, all LLMs achieved an NPV >98% comparable to the human benchmark (99.0%; 95% CI 97.9%–99.8%). However, poor specificity resulted in substantially lower model PPVs (range 70.0%–77.5%) compared to humans (96.6%; 95% CI 92.8%–99.3%) (eFigure 10, eTable 3).

### Secondary Outcomes

For classification of the primary tumor diagnosis, human accuracy was 95.2% (95% CI 93.4%–97.0%). Only gpt-oss:120b was non-inferior (94.0%; 95% CI 91.3%–96.7%). Accuracy varied by tumor type, with models generally performing similarly to humans (eFigure 11), except for qwq:32b and qwen3:32b, which performed poorly. For the classification of radiological tumor absence, human accuracy was 87.5% (95% CI 84.8%–90.2%). All LLMs were inferior to human classification. Across all tasks only gpt-oss:120b was non-inferior (Figure 3, eTable 4).

**Figure 3:**
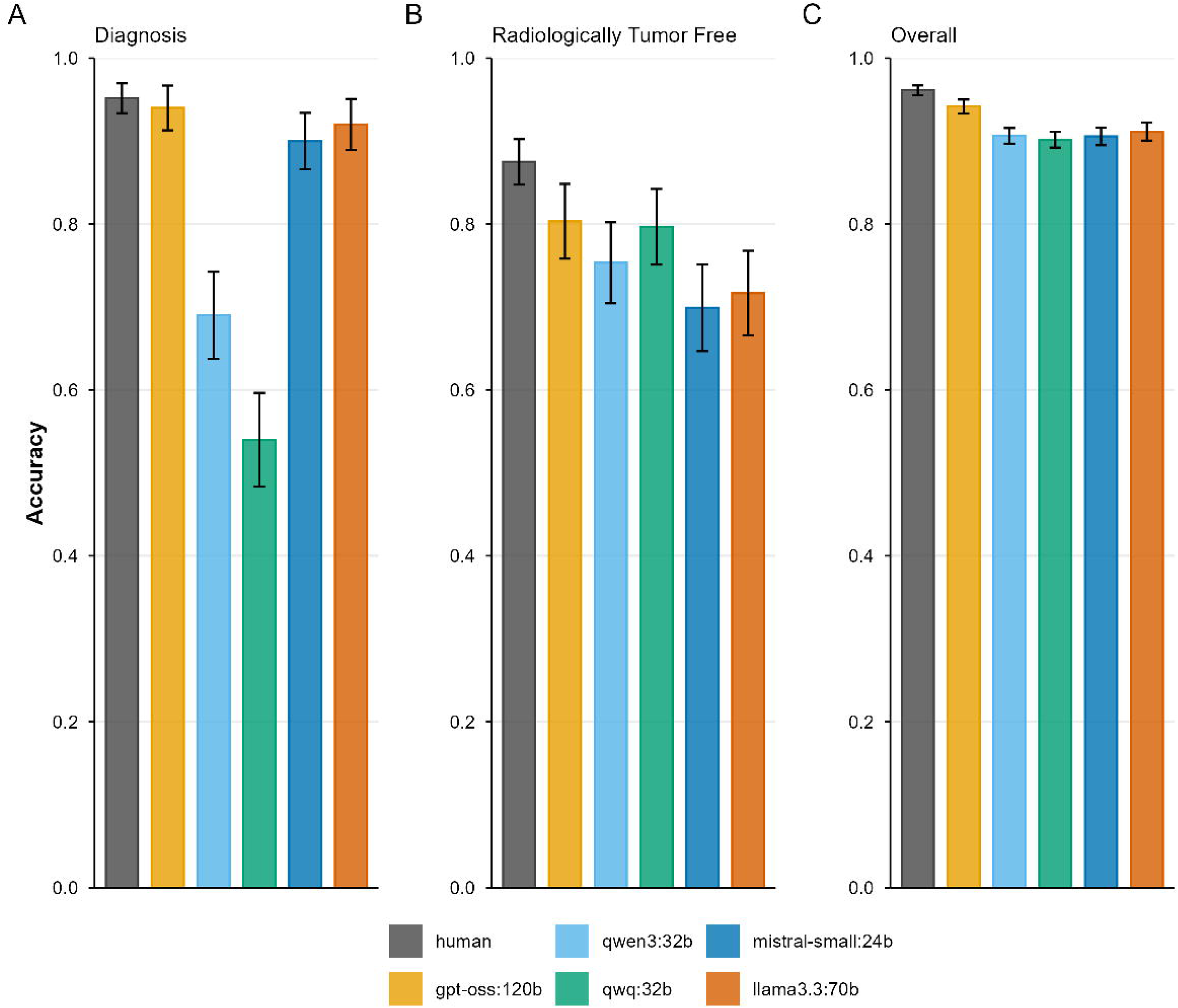
Plots show pooled accuracy and 95% confidence intervals (CIs) comparing humans and LLMs. **(A)** Multi-class classification of primary tumor diagnosis. **(B)** Binary classification of radiological absence of tumor (yes vs. no). **(C)** Overall accuracy pooled across all primary (Figure 1) and secondary (Figure 3A, 3B) endpoint tasks.

Mean human extraction time per report was 120s (range 60–240s). LLM processing times per report were 11s (mistral-small:24b), 33s (llama3.3:70b), 45s (gpt-oss:120b), 47s (qwq:32b), and 63s (qwen3:32b).

### Sensitivity Analyses

A Bayesian sensitivity analysis yielded results consistent with the frequentist analysis (eFigures 12–14). In a further sensitivity analysis to assess ground truth reliability, blinded senior oncologists reviewed all discrepancies between the ground truth and gpt-oss:120b for the primary outcomes. While the original ground truth was confirmed in most cases (Metastasis: 48/64; Response: 50/64), the changes were sufficient for gpt-oss:120b to achieve non-inferiority for treatment response when evaluated against the revised ground truth (eFigure 15, eTable 5).

## Discussion

In this study, we show that locally hosted LLMs can achieve human-level accuracy for explicit data extraction tasks in German radiology reports but underperform humans when interpretive clinical reasoning is required. Specifically, while all models identified metastatic sites with high accuracy, none achieved non-inferiority for the complex task of treatment response assessment. Similar results have been observed in different datasets in different languages ^2,24^.

This performance decrease can likely be explained in part by data quality: the radiology reports often contained ambiguous language or omitted definitive conclusions, making ground truth establishment challenging. This ambiguity mirrors clinical reality, where determining whether imaging findings truly reflect a response is often time-consuming and requires interdisciplinary discussion. Human interrater agreement decreased for classification of treatment response and radiological tumor absence, suggesting that the information required for a definitive classification was sometimes absent from the report itself. Integrating longitudinal data, such as previous imaging reports, clinical notes, and treatment changes might improve accuracy but would significantly increase pipeline complexity.

We observed unexpected variability in model performance across tasks: qwen3:32B and qwq:32b performed robustly for metastasis classification but poorly for primary diagnosis classification, even after prompt optimization. This inconsistency suggests that performance cannot easily be extrapolated from one task to another; models should be benchmarked specifically for each intended use case before deployment.

We also found that implementation requires iterative prompt optimization and pipeline adaptation. We observed substantial performance gains by modifying our pipeline to allow models to generate a free-text reasoning step before outputting the structured JSON format, except for gpt-oss:120b, which performed best with a single prompt strategy.

Our study has several strengths. We adhered to rigorous clinical research standards, including preregistration of the study, extraction in duplicate, and adjudication by a senior oncologist. Furthermore, our conclusions are robust to multiplicity; when applying a Bonferroni correction for ten comparisons (five models across two primary endpoints), the non-inferiority (p < 0.001) of all models for metastasis status would remain unchanged.

Our study has limitations. The single-center, German-language setting limits generalizability to other languages and reporting styles. Furthermore, we were not able to compare the performance of LLMs to more established methods in natural language processing, such as bidirectional encoder representation from transformers, which have been successfully implemented for routine oncology data ^2,25^. This was unfeasible because we did not have the resources to annotate enough data for fine-tuning models for each classification task ^2^. The availability of only a single radiology report, without additional information or clinical context (e.g., prior imaging or the patient’s clinical status), represents an artificial scenario that may have adversely affected the performance of both human coders and large language models. Finally, we restricted our evaluation to open-source, locally hosted models to prioritize data privacy. It is likely that larger commercial models would achieve higher accuracy. Finally, because the ground truth relied on human consensus, it inherently favors human performance. Our sensitivity analysis revealed that in ca. 20% of adjudicated discrepancies between the ground truth and gpt-oss:120b, the original ground truth was deemed incorrect. This finding highlights a fundamental challenge in benchmarking LLMs on real-world clinical data: establishing a definitive gold standard is inherently difficult due to the complexity and ambiguity of medical reports.

Despite these limitations, LLMs could, when used carefully, improve research efficiency. For example, determination of progression-free survival, a critical endpoint in oncology, typically requires labor-intensive manual review of longitudinal imaging to identify the date of first progression. Automating the extraction of progression events or new metastases could significantly accelerate this process. While our findings suggest that current open-source solutions are not yet robust enough for fully automated implementation, the rapid pace of advancement indicates that future models may increasingly become viable for these complex applications.

## Conclusions

Locally hosted LLMs likely provide a non-inferior alternative to human extraction for more explicit clinical data elements like metastasis location. However, for more interpretive tasks involving ambiguity or longitudinal context, current open-source models remain inferior to human extraction. Successful integration of these tools into clinical research pipelines requires task-specific validation and careful consideration of model choice.

## Supporting information

TRIPOD+LLM-checklist

## Data Availability

We are unable to share the original clinical reports because they contain protected patient data; however, all analysis code and code used for prompting is available on GitHub.

https://github.com/scjohannes/llm-med-extraction

## Abbreviations

CDWH: Clinical Data Warehouse
CI: Confidence Interval
CNS: Central Nervous System
CT: Computed Tomography
MRI: Magnetic Resonance Imaging
PET: Positron Emission Tomography
GPU: Graphics Processing Unit
LLM: Large Language Model
NPV: Negative Predictive Value
PPV: Positive Predictive Value
RANO: Response Assessment in Neuro-Oncology
RECIST: Response Evaluation Criteria in Solid Tumors
REDCap: Research Electronic Data Capture
y: year
pp: percentage points
SD: Standard Deviation

## Declarations

## Acknowledgements

We thank Noah Zanolari and Anne Michel for supporting the data extraction.

## Authors’ contributions

1. Conceptualization: JMS, BK
2. Data Curation: JMS, LPP
3. Formal Analysis: JMS, LPP
4. Funding Acquisition: -
5. Investigation: LPP, JMS, CMS, FL, JP, BT, DH, AD, AMS, BK
6. Methodology: JMS
7. Project Administration: JMS
8. Resources: MD
9. Software: JMS, MD
10. Supervision: MB, BK
11. Validation: BK, AMS
12. Visualization: JMS, LPP
13. Writing – Original Draft: JMS, LPP
14. Writing – Review & Editing: all authors

## Availability of data and materials

We are unable to share the original clinical reports because they contain protected patient data; however, all analysis code and code used for prompting is available on GitHub ^16^.

## Ethics approval and consent to participate

Ethical approval was obtained from the Ethics Committee Northwestern Switzerland (BASEC 2025–01036). The study design and analyses were preregistered on OSF^17^.

## Consent for publication

N/A

