## Supplementary Appendix for "Data Extraction from Oncology Imaging Reports by Large Language Models: A Comparative Accuracy Study"

Supplementary Materials

2025-12-29

### eFigures

#### eFigure 1

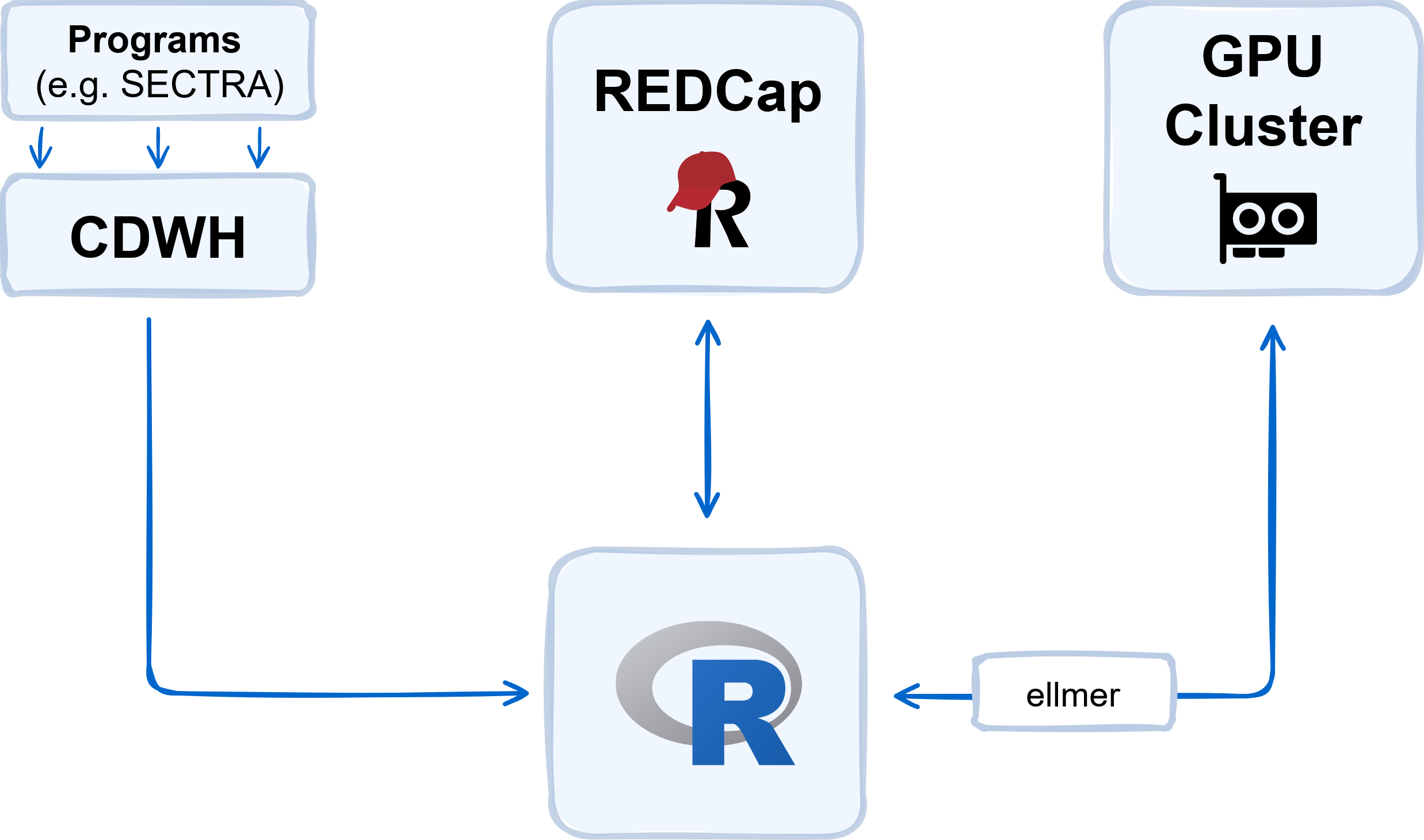

eFigure 1: This figure shows the flow of data in the study. The unstructured radiology reports and metadata are pulled from the clinical data warehouse (CDWH) using a database interface in R and uploaded to the study database hosted on a local REDCap instance. The imaging reports are processed by Large Language Models (LLMs) on a local GPU cluster. For communication between R and the LLMs we use the ellmer package. Abbreviations: CDWH, clinical data warehouse; GPU, graphics processing unit.

#### eFigure 2

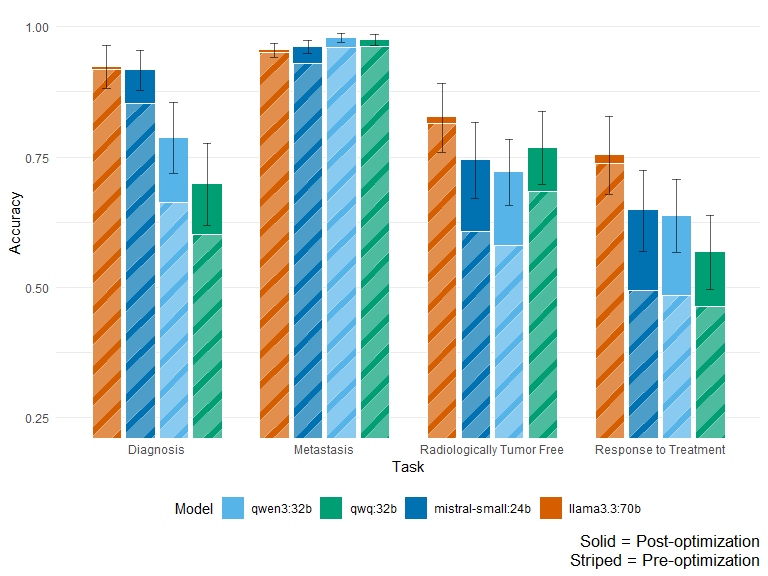

eFigure 2: The plot shows the accuracy before and after prompt optimization for each model for each task in the prompt optimization data (n = 100). The striped bars show the pre-optimization accuracy, the solid bars show the post-optimization accuracy. Error bars represent 95% confidence intervals calculated using cluster-robust standard errors for post-optimization accuracy. We gained access to gpt-oss:120b after prompt optimization, hence it is not included in this analysis, as no pre-optimization data exists.

#### eFigure 3

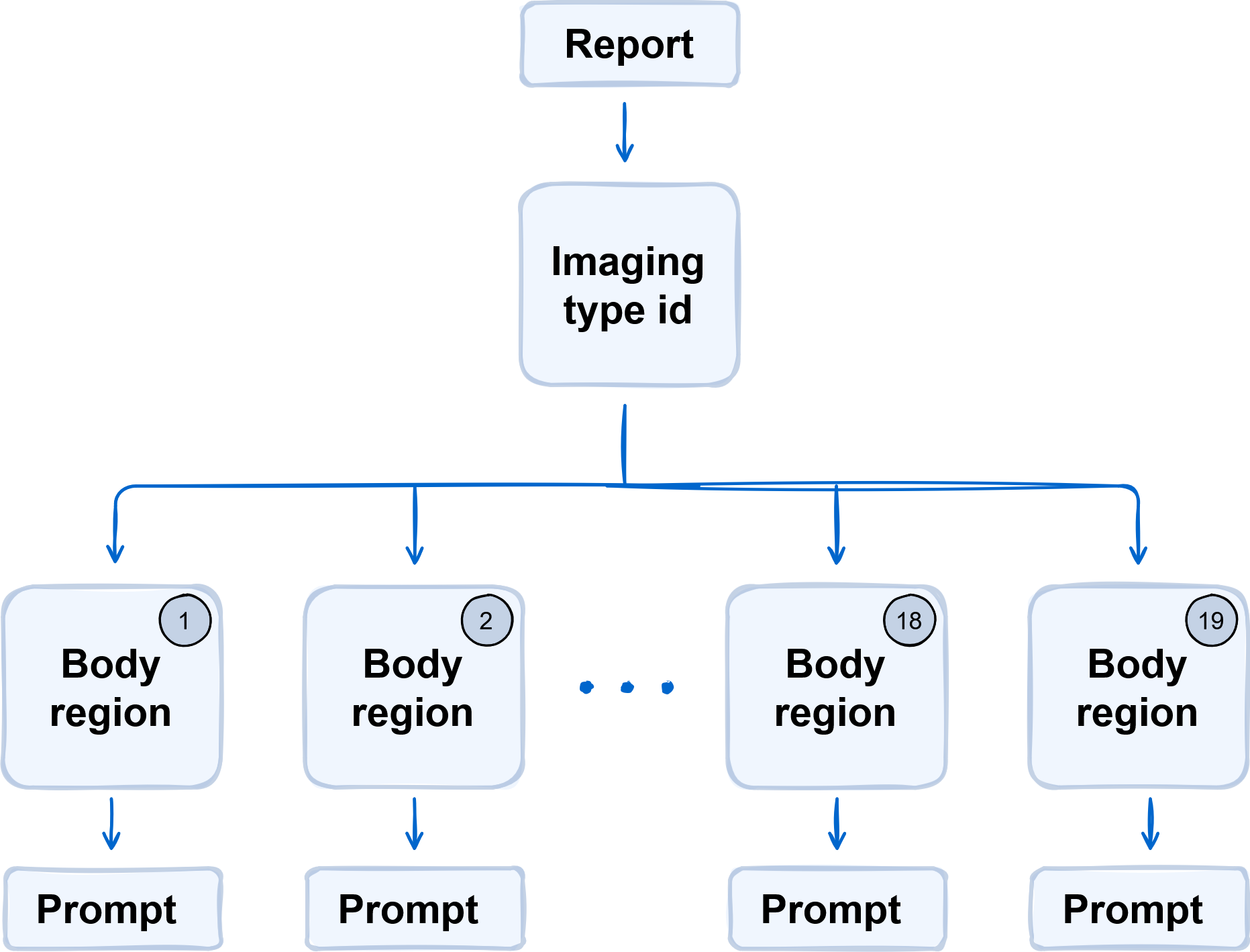

eFigure 3: This figure shows how prompts for classification of metastasis are adapted to the imaging report. Each report has an ‘imaging type’ identifier in its metadata. Based on this identifier, the report is automatically classified as pertaining to one of 19 body regions and the corresponding prompt is selected. The prompts used for metastasis classification are shown in eMethods.

#### eFigure 4

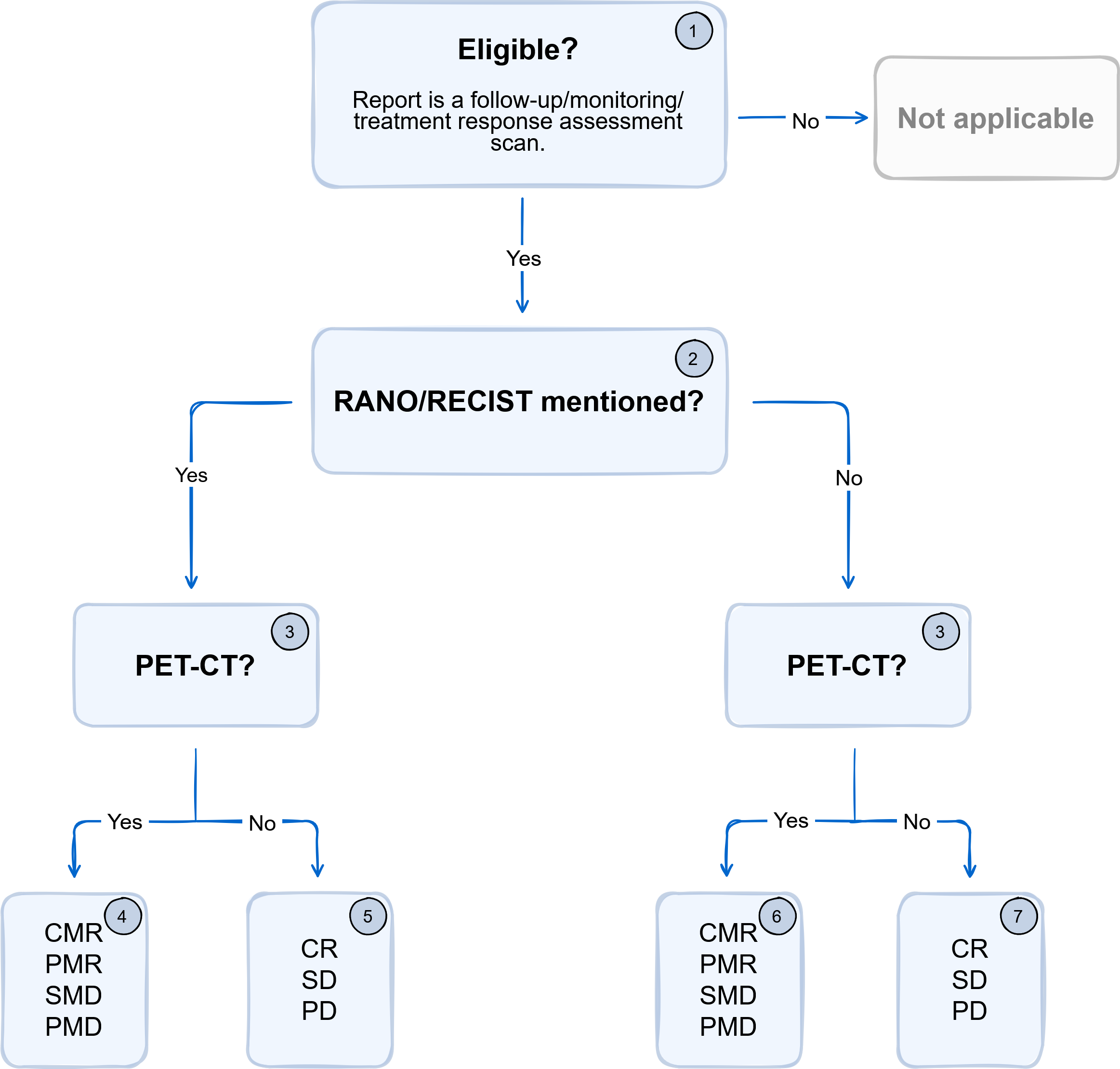

eFigure 4: This figure shows how prompts for classification of response to treatment are chained. (1) A prompt to check the imaging report for being relevant to response to treatment is used. (2) The reports classified as eligible (in-scope) are categorized into reports mentioning RANO or RECIST using regular expressions. (3) The prompts for data extraction are selected according to whether the imaging report was a PET-scan, based on the report metadata. (4-7) The prompts used for data extraction are shown in the eMethods.

#### eFigure 5

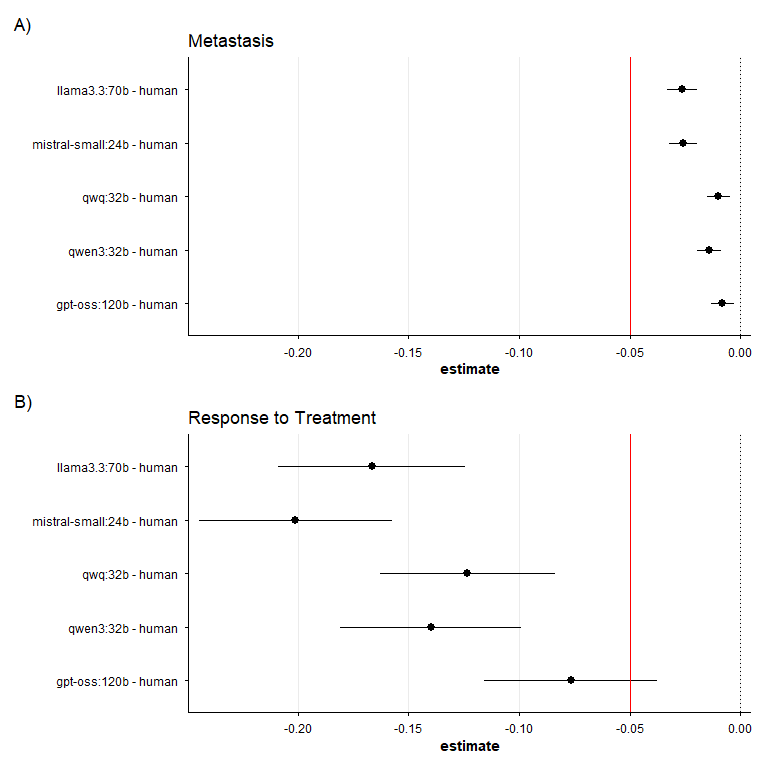

eFigure 5: Difference in the accuracy of models compared to human extractors for the primary endpoints: Metastasis Detection (A) and Response to Treatment Classification (B). Point estimates with 90% confidence intervals are shown. The red solid line indicates the pre-specified equivalence margin of -5%. If the lower bound of the confidence interval is above this margin, the model is considered non-inferior to human extractors.

#### eFigure 6

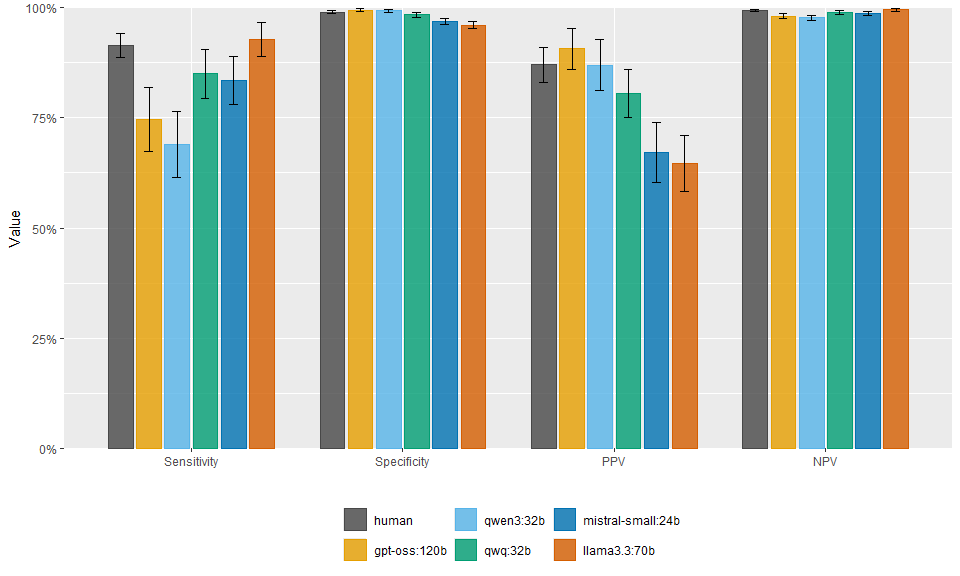

eFigure 6: Sensitivity, specificity, positive predictive value (PPV), and negative predictive value (NPV) for metastasis detection by human extractors and models. Point estimates with 95% confidence intervals are shown.

#### eFigure 7

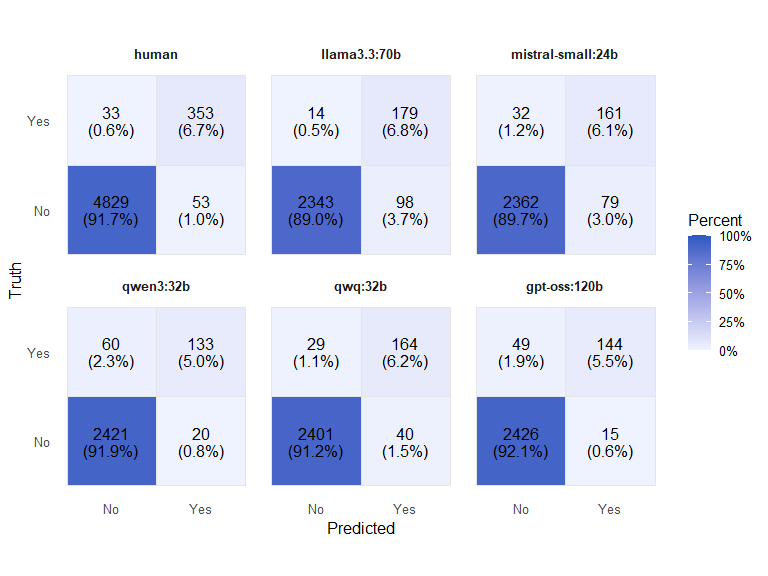

eFigure 7: Confusion matrices for human extractors and models for metastasis classification. As humans extracted in duplicate, the confusion matrix for humans is based on both extractions.

#### eFigure 8

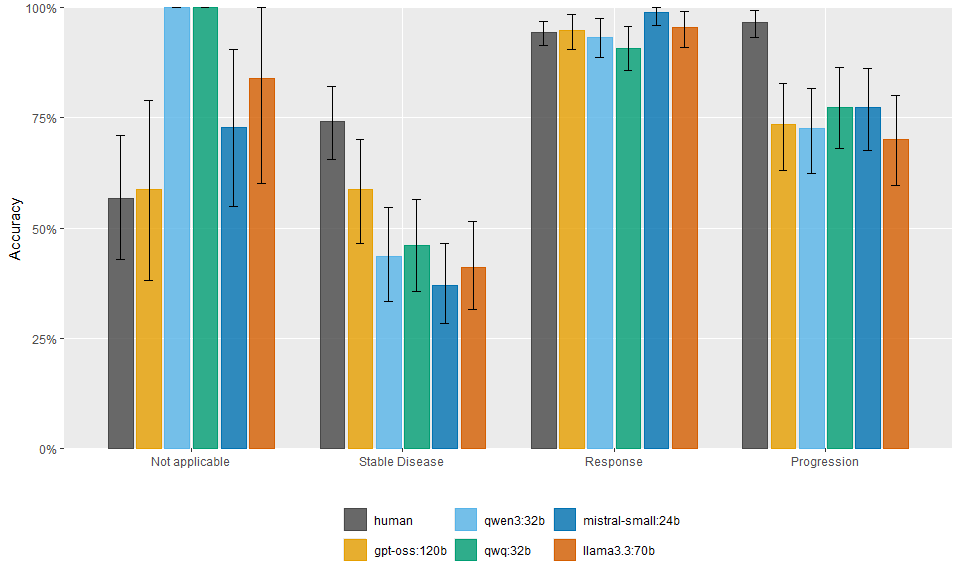

eFigure 8: Accuracy of human extractors and models for response to treatment classification. Point estimates with bootstrapped 95% confidence intervals (1000 repetitions) are shown. The sample size of PET-scans was too small to accurately estimate accuracy for partial response and complete response separately; these two categories were therefore merged into a single ‘response’ category.

#### eFigure 9

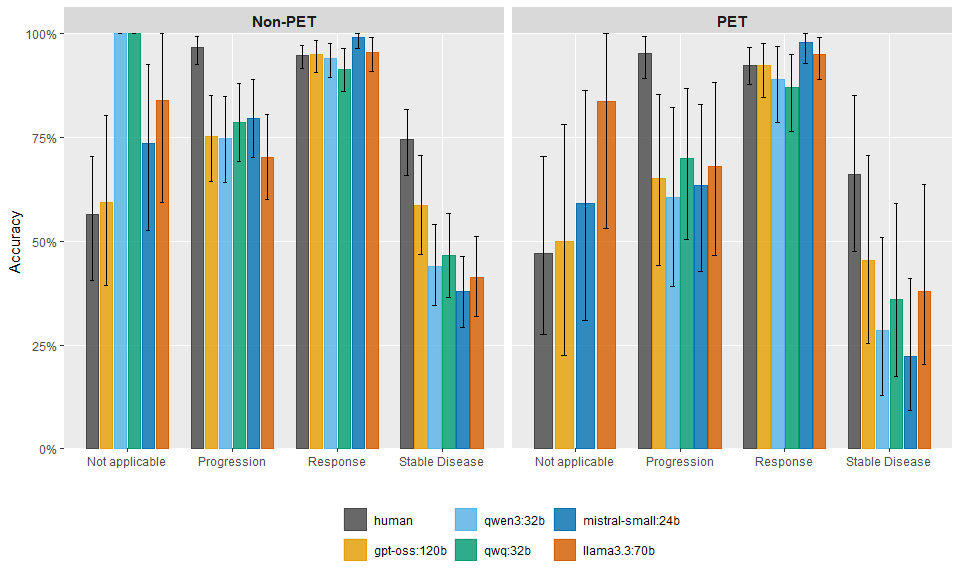

eFigure 9: Accuracy of human extractors and models for response to treatment classification, stratified by imaging modality (CT/MRI vs. PET-scan) and classification. Point estimates with bootstrapped 95% confidence intervals (1000 repetitions) are shown. The sample size of PET-scans was too small to accurately estimate accuracy for partial response and complete response separately; these two categories were therefore merged into a single ‘response’ category.

#### eFigure 10

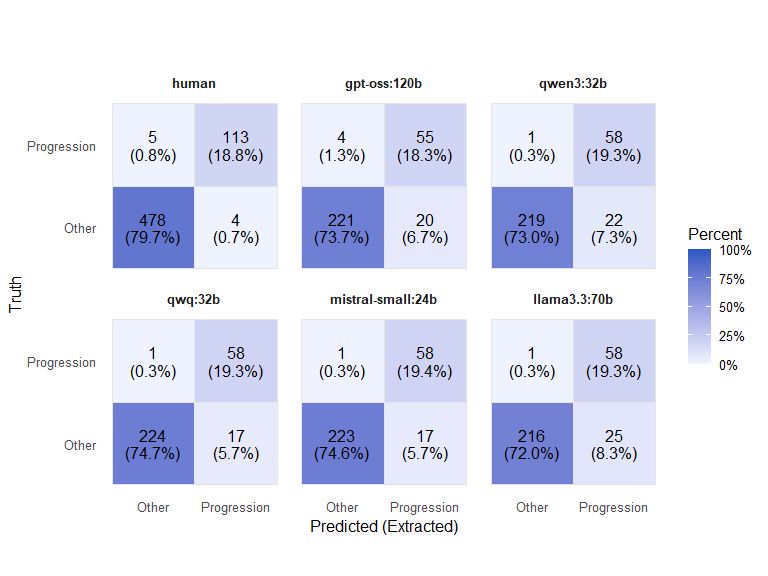

eFigure 10: Confusion matrices for humans and models for response to treatment classification, when response to treatment is dichotomized into progression vs other. As humans extracted in duplicate, the confusion matrix for humans is based on both extractions.

#### eFigure 11

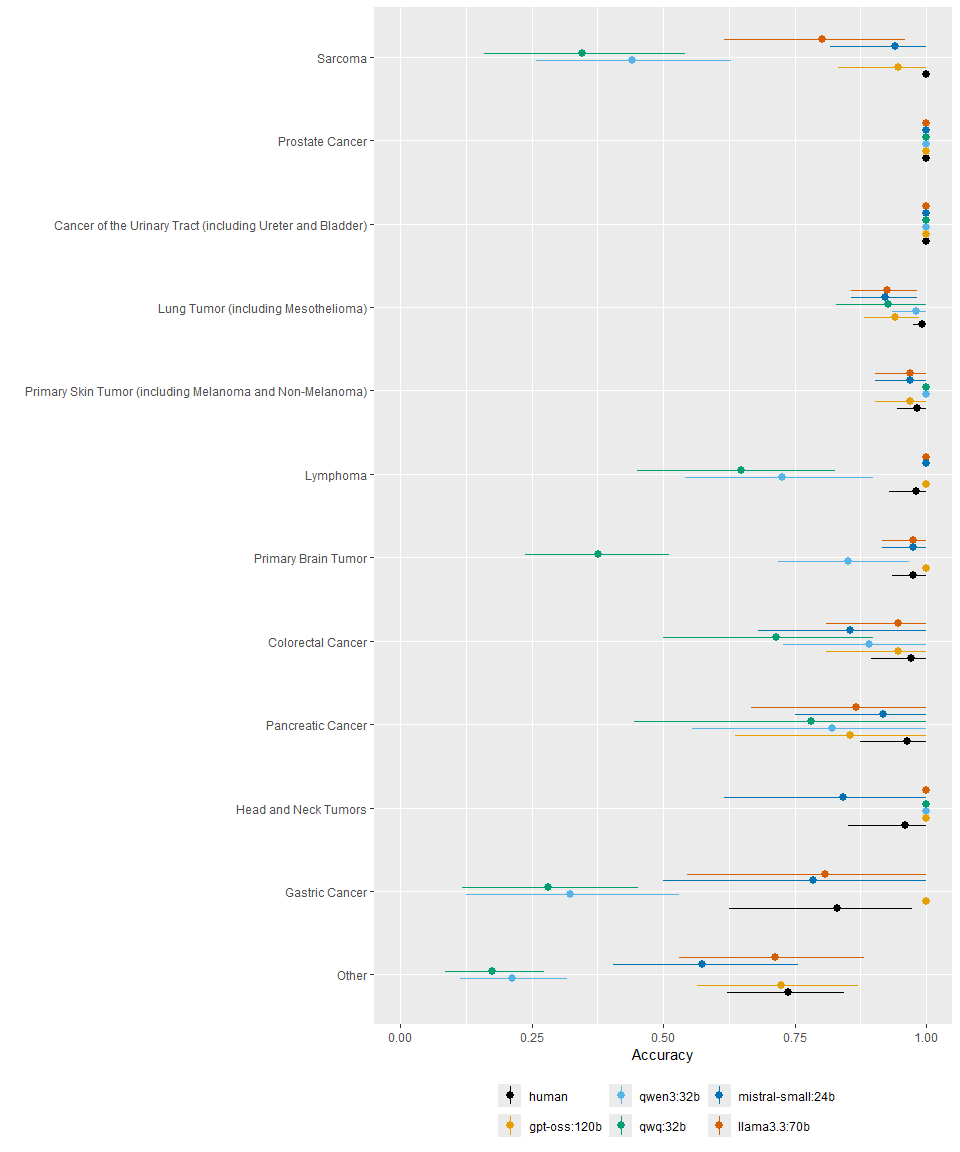

eFigure 11: This plot shows the accuracy with 95% confidence intervals of human extractors and model extraction compared to the ground truth, stratified by diagnosis. Diagnoses with fewer than 50 extractions were grouped into ‘Other’. Confidence intervals were calculated using cluster bootstrapping resampling (1000 samples).

#### eFigure 12

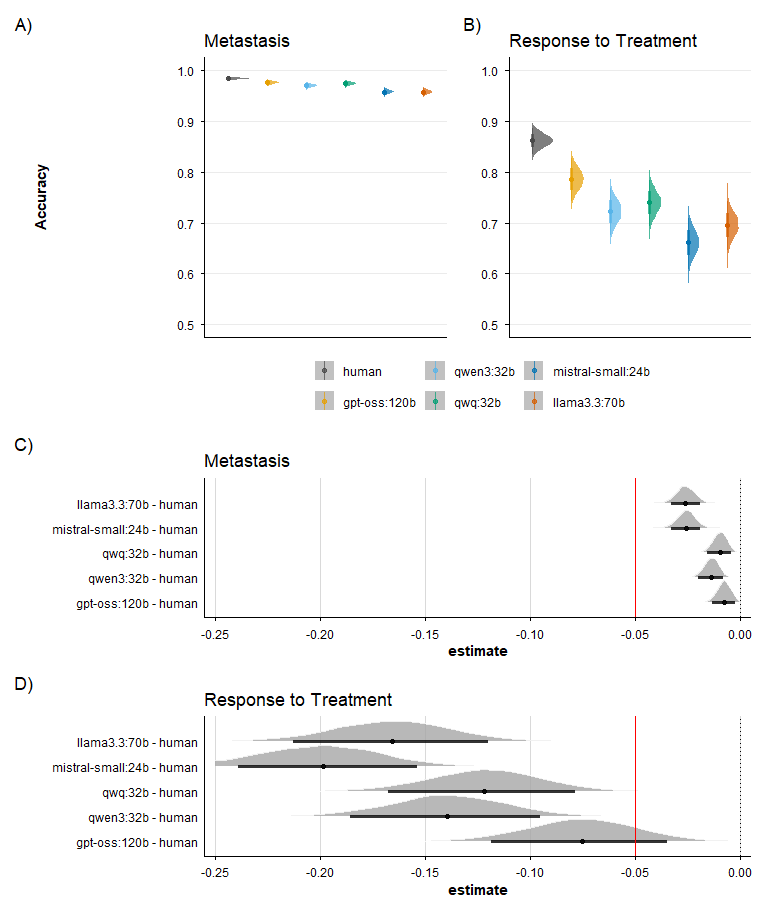

eFigure 12: Bayesian reanalysis of the accuracy for classification of metastasis and response to treatment. Top panels (A, B) show the posterior distributions of accuracy (probability of correct response) for humans and each model across the two primary tasks with 66% and 95% credible intervals. Bottom panels (C, D) show the posterior distributions of the contrasts (model accuracy - human accuracy) and 90% credible intervals for each model compared to humans, with the non-inferiority margin indicated by the solid red line at -0.05.

#### eFigure 13

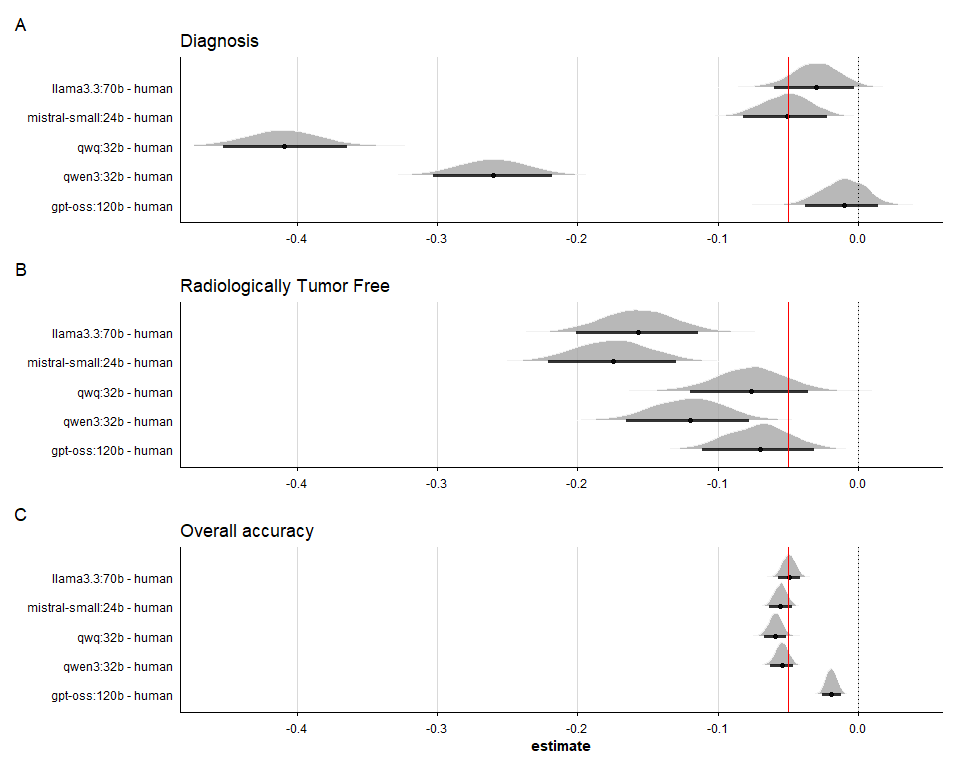

eFigure 13: Bayesian reanalysis of contrasts (model accuracy - human accuracy) for secondary endpoints (diagnosis, radiologically tumor free, overall accuracy); posterior distributions with 90% credible intervals shown.

#### eFigure 14

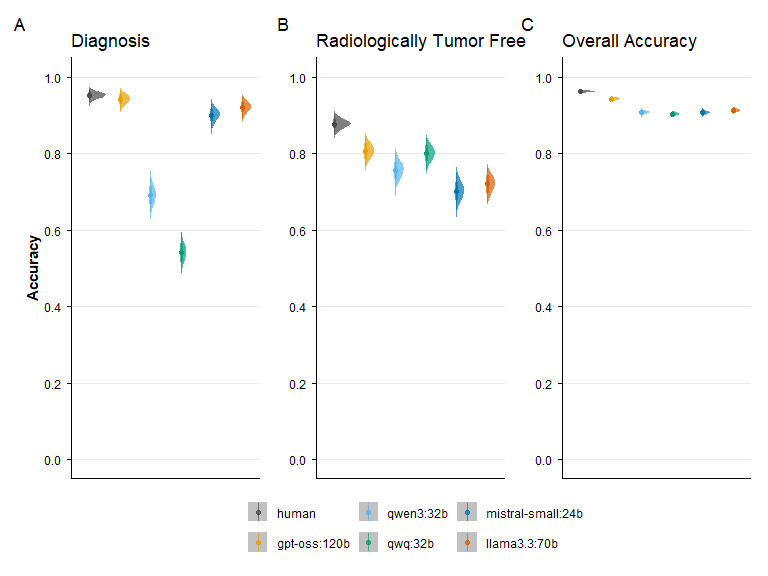

eFigure 14: Bayesian reanalysis of accuracy for secondary endpoints (diagnosis, radiologically tumor free, overall accuracy); posterior distributions with 66% and 95% credible intervals shown.

#### eFigure 15

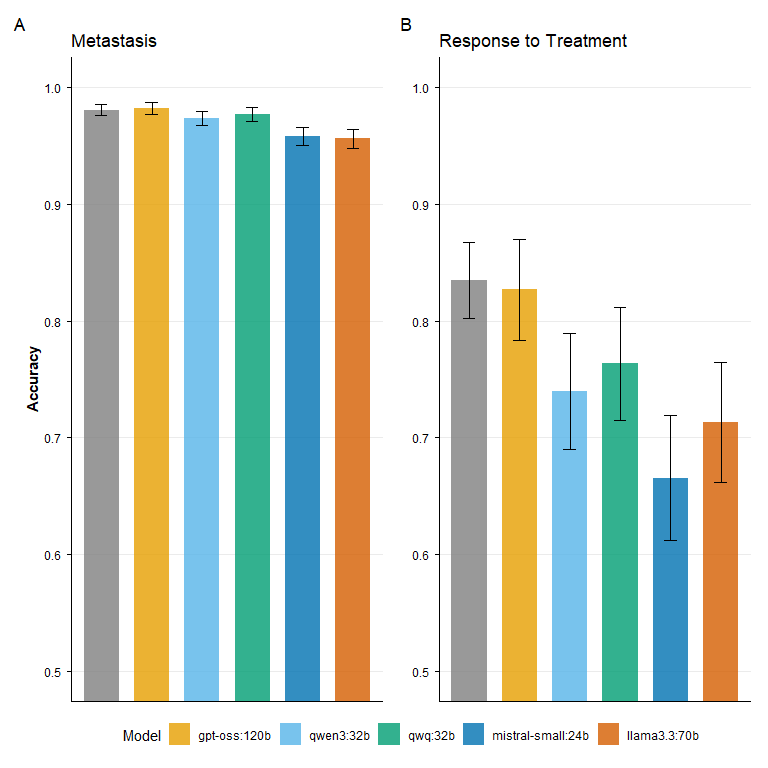

eFigure 15: Sensitivity analysis using readjudicated ground truth for the primary endpoints. For cases where gpt-oss:120b disagreed with the original ground truth, two blinded senior oncologists reviewed and readjudicated the cases. (A) Metastasis detection accuracy. (B) Response to treatment classification accuracy. Point estimates with 95% confidence intervals calculated using cluster-robust standard errors are shown.

### eTables

#### eTable 1

|  | Sensitivity (95% CI) | Specificity (95% CI) | PPV (95% CI) | NPV (95% CI) |
| --- | --- | --- | --- | --- |
| human | 91.5% (88.7–94.2%) | 98.9% (98.6–99.2%) | 86.9% (83.1–90.8%) | 99.3% (99.1–99.6%) |
| gpt-oss:120b | 74.6% (67.3–81.9%) | 99.4% (99.1–99.7%) | 90.6% (86.0–95.1%) | 98.0% (97.4–98.7%) |
| qwen3:32b | 68.9% (61.5–76.3%) | 99.2% (98.8–99.6%) | 86.9% (81.1–92.7%) | 97.6% (97.0–98.2%) |
| qwq:32b | 85.0% (79.4–90.5%) | 98.4% (97.8–98.9%) | 80.4% (74.9–85.8%) | 98.8% (98.3–99.3%) |
| mistral-small:24b | 83.4% (78.1–88.8%) | 96.8% (96.1–97.5%) | 67.1% (60.4–73.8%) | 98.7% (98.2–99.1%) |
| llama3.3:70b | 92.7% (88.9–96.6%) | 96.0% (95.1–96.8%) | 64.6% (58.3–70.9%) | 99.4% (99.1–99.7%) |

Table 1: Sensitivity, specificity, positive predictive value (PPV), and negative predictive value (NPV) for metastasis classification by human extractors and models.

#### eTable 2

|  | Accuracy % (95% CI) |
| --- | --- |
| Not applicable | |
| human | 56.7% (42.9%–71.1%) |
| gpt-oss:120b | 58.7% (38.1%–78.9%) |
| qwen3:32b | 100.0% (100.0%–100.0%) |
| qwq:32b | 100.0% (100.0%–100.0%) |
| mistral-small:24b | 72.9% (55.0%–90.5%) |
| llama3.3:70b | 84.0% (60.0%–100.0%) |
| Progression | |
| human | 96.7% (93.1%–99.3%) |
| gpt-oss:120b | 73.4% (63.0%–82.7%) |
| qwen3:32b | 72.5% (62.3%–81.6%) |
| qwq:32b | 77.4% (67.9%–86.3%) |
| mistral-small:24b | 77.4% (67.6%–86.2%) |
| llama3.3:70b | 70.0% (59.7%–80.0%) |
| Response | |
| human | 94.3% (91.4%–96.8%) |
| gpt-oss:120b | 94.7% (90.5%–98.4%) |
| qwen3:32b | 93.2% (88.6%–97.5%) |
| qwq:32b | 90.8% (85.7%–95.6%) |
| mistral-small:24b | 98.8% (95.9%–100.0%) |
| llama3.3:70b | 95.4% (90.8%–99.0%) |
| Stable Disease | |
| human | 74.1% (65.5%–82.1%) |
| gpt-oss:120b | 58.6% (46.5%–70.0%) |
| qwen3:32b | 43.6% (33.3%–54.8%) |
| qwq:32b | 46.1% (35.6%–56.4%) |
| mistral-small:24b | 37.1% (28.4%–46.5%) |
| llama3.3:70b | 41.1% (31.5%–51.6%) |

eTable 2: Accuracy with 95% bootstrap confidence intervals (1000 repetitions) for response to treatment classification by human extractors and models. For PET‑scans partial and complete responses were combined into a single “response” category.

#### eTable 3

|  | Sensitivity (95% CI) | Specificity (95% CI) | PPV (95% CI) | NPV (95% CI) |
| --- | --- | --- | --- | --- |
| human | 95.6% (91.8–98.6%) | 83.6% (80.4–86.6%) | 96.6% (92.8–99.3%) | 99.0% (97.9–99.8%) |
| gpt-oss:120b | 93.3% (86.0–98.6%) | 74.8% (69.3–80.1%) | 73.5% (63.5–83.3%) | 98.2% (96.3–99.6%) |
| qwen3:32b | 98.3% (94.6–100.0%) | 65.5% (59.7–71.5%) | 72.7% (63.0–82.9%) | 99.5% (98.6–100.0%) |
| qwq:32b | 98.3% (94.0–100.0%) | 67.6% (61.7–73.6%) | 77.5% (66.7–87.0%) | 99.6% (98.6–100.0%) |
| mistral-small:24b | 98.3% (94.7–100.0%) | 58.0% (52.0–64.0%) | 77.5% (67.5–87.0%) | 99.5% (98.6–100.0%) |
| llama3.3:70b | 98.3% (94.6–100.0%) | 62.3% (55.7–68.3%) | 70.0% (60.7–79.8%) | 99.5% (98.5–100.0%) |

eTable 3: Sensitivity, specificity, positive predictive value (PPV), and negative predictive value (NPV) for response to treatment, when dichotomized into progression vs other, by human extractors and models.

#### eTable 4

|  |  | **Accuracy**  % (95% CI) | **Difference** **from Human** Percentage Points (90% CI) | **P-value**  Noninferiority |
| --- | --- | --- | --- | --- |
|  | Diagnosis | | | |
|  | human | 95.2% (93.4%–97.0%) |  |  |
|  | gpt-oss:120b | 94.0% (91.3%–96.7%) | -1.2 (-3.3 to 1.0) | 0.002 |
|  | qwen3:32b | 69.0% (63.8%–74.2%) | -26.2 (-30.6 to -21.8) | >0.999 |
|  | qwq:32b | 54.0% (48.4%–59.6%) | -41.2 (-45.9 to -36.4) | >0.999 |
|  | mistral-small:24b | 90.0% (86.6%–93.4%) | -5.2 (-7.9 to -2.5) | 0.540 |
|  | llama3.3:70b | 92.0% (88.9%–95.1%) | -3.2 (-5.4 to -0.9) | 0.091 |
|  | Radiologically Tumor Free | | | |
|  | human | 87.5% (84.8%–90.2%) |  |  |
|  | gpt-oss:120b | 80.3% (75.8%–84.8%) | -7.2 (-10.6 to -3.7) | 0.848 |
|  | qwen3:32b | 75.3% (70.4%–80.2%) | -12.2 (-15.8 to -8.5) | >0.999 |
|  | qwq:32b | 79.7% (75.1%–84.2%) | -7.8 (-11.3 to -4.3) | 0.909 |
|  | mistral-small:24b | 69.9% (64.7%–75.1%) | -17.6 (-21.7 to -13.6) | >0.999 |
|  | llama3.3:70b | 71.7% (66.6%–76.8%) | -15.8 (-19.8 to -11.9) | >0.999 |
|  | Overall | | | |
|  | human | 96.1% (95.5%–96.7%) |  |  |
|  | gpt-oss:120b | 94.2% (93.3%–95.0%) | -2.0 (-2.6 to -1.3) | <0.001 |
|  | qwen3:32b | 90.6% (89.7%–91.6%) | -5.5 (-6.3 to -4.7) | 0.843 |
|  | qwq:32b | 90.2% (89.2%–91.1%) | -5.9 (-6.7 to -5.1) | 0.975 |
|  | mistral-small:24b | 90.6% (89.5%–91.6%) | -5.6 (-6.4 to -4.7) | 0.857 |
|  | llama3.3:70b | 91.1% (90.1%–92.2%) | -5.0 (-5.9 to -4.1) | 0.485 |
| eTable 4: Accuracy with 95% confidence intervals for diagnosis, radiologically tumor free, and across all tasks, by human extractors and LLMs, along with comparisons to human performance with 90% confidence intervals. | | | | |

#### eTable 5

|  | **Accuracy** | **Difference from Human** | **P-value** |
| --- | --- | --- | --- |
|  | % (95% CI) | Percentage Points (90% CI) | Noninferiority |
| Metastasis | | | |
| human | 98.0% (97.5%–98.5%) | [Reference] |  |
| gpt-oss:120b | 98.2% (97.6%–98.7%) | 0.2 (-0.4 to 0.7) | <0.001 |
| qwen3:32b | 97.3% (96.7%–98.0%) | -0.7 (-1.3 to -0.1) | <0.001 |
| qwq:32b | 97.7% (97.1%–98.3%) | -0.3 (-0.9 to 0.2) | <0.001 |
| mistral-small:24b | 95.8% (95.0%–96.6%) | -2.2 (-2.9 to -1.6) | <0.001 |
| llama3.3:70b | 95.6% (94.8%–96.4%) | -2.4 (-3.1 to -1.8) | <0.001 |
| Response to Treatment | | | |
| human | 83.5% (80.3%–86.7%) | [Reference] |  |
| gpt-oss:120b | 82.7% (78.4%–87.0%) | -0.8 (-4.9 to 3.2) | 0.046 |
| qwen3:32b | 74.0% (69.0%–79.0%) | -9.5 (-13.8 to -5.2) | 0.959 |
| qwq:32b | 76.3% (71.5%–81.2%) | -7.2 (-11.3 to -3.0) | 0.807 |
| mistral-small:24b | 66.6% (61.2%–71.9%) | -16.9 (-21.4 to -12.5) | >0.999 |
| llama3.3:70b | 71.3% (66.2%–76.5%) | -12.2 (-16.6 to -7.8) | 0.996 |

eTable 5: Sensitivity analysis using readjudicated ground truth for the primary endpoints. Accuracy with 95% confidence intervals, differences from human extractors with 90% confidence intervals, and noninferiority p-values are shown. Cases where gpt-oss:120b disagreed with the original ground truth were reviewed and readjudicated by two blinded senior oncologists.

### eMethods

#### Prompt and Pipeline Optimization

##### Pipeline Adaptation

We refined prompts using a prompt-optimization set ($n=100$). Initial evaluations showed that requesting immediate structured output frequently resulted in incorrect responses, where the model’s textual justification contradicted the extracted JSON value. To address this, we adjusted the pipeline to allow a model to first answer in free-text, and in a second step in the same chat output its response as a structured JSON. This approach led to improvements in performance across most models.

##### Model-Specific Configuration

We gained access to gpt-oss:120b after the initial prompt optimization phase. Evaluation on the optimization set indicated that, unlike the other models, the multi-step reasoning pipeline worsened performance for this specific model. Consequently, we utilized a single integrated prompt for gpt-oss:120b that requested reasoning and JSON output simultaneously.

##### Task-Specific Scope Assessment

For the *response to treatment* and *radiologically tumor free* tasks, we identified a specific issue where models frequently classified relevant reports as “Not Applicable” (out of scope) when tasked with assessing scope and classification simultaneously. To mitigate this issue of incorrectly excluding valid reports, we split the pipeline into distinct stages (see eFigure 3). First, a dedicated prompt asked the model to determine if the report was within the scope of the clinical question. If affirmed, subsequent prompts were triggered to perform the specific data extraction and classification.

##### Ground Truth Validation

To ensure the integrity of the study data, a senior oncologist (BK), blinded to the LLM predictions, reviewed all 43 reports that had been labeled as “out of scope” by the human abstractors. This review determined that 13 (30.2%) of these reports were, in fact, in scope and contained valid information for response assessment. These reports were subsequently relabeled and included in the final analysis.

#### Extraction Guidelines for Humans

##### Extraction of Primary Tumor Diagnosis

- Extract the location (organ) of the primary tumor from the imaging exam report.
- Often the primary location is directly mentioned in the ‘Anamnese’ section of the report. The primary location of the tumor and its name in the ‘Anamnese’ section can be treated as the same thing.
- It is possible that the primary location of the tumor is unclear or not mentioned, but we know location of metastases. In that case choose ‘Unclear’. • Do not use the location of metastasis to make a guess about the location of the primary tumor.
- Choose ‘Not Applicable’ if the the clinical context/findings clearly indicate the scan was performed for reasons unrelated to the cancer.
- If the patient has two tumors simultaneously, choose the tumor that is mentioned first in the report.

##### Extraction of Metastasis Presence

You will be given a report of the imaging exam of the of a patient with cancer. You are asked to extract whether this patient presently has metastasis in the imaged are as mentioned in the report.

Please answer for each questions displayed below:

**Interpreting PET-CT Findings:**

- Pay close attention to descriptions of lesions, particularly those identified as ‘hypermetabolic’, ‘FDG-avid’, having ‘increased uptake’, or showing a specific ‘SUV’ (Standardized Uptake Value). These indicate metabolic activity often associated with cancer but require careful interpretation within the report’s context.
- Radiologists may use terms like ‘lesion’, ‘nodule’, ‘mass’, ‘uptake’, or ‘activity’ instead of explicitly stating ‘metastasis’. Your task is to determine if the description and context imply metastatic disease.

**Interpreting CT and MRI-Findings:**

- Identify if the radiologist explicitly labels any finding as ‘Metastase’, ‘metastasenverdächtig’, or similar.
- Assess if the radiologist describes lesions/nodules/masses with characteristics and then explicitly states they are suspicious for, concerning for, consistent with, likely, or typical of metastasis.
- Note if the radiologist offers a differential diagnosis; the likelihood assigned to metastasis is key (see TRUE/FALSE criteria).

**Answering TRUE/FALSE:**

- Answer with TRUE if there is:
- A clear mention of metastasis in the specific location.
- A description of a lesion where the radiologist expresses a strong suspicion or high likelihood of it representing metastasis (e.g., ‘metastasenverdächtig’, ‘concerning for metastatic deposit’, ‘vereinbar mit Metastasierung’, ‘wahrscheinliche Metastasen’).
- Answer with FALSE if:
  - Metastasis is mentioned but considered less likely than other possibilities (e.g., listed lower in the differential diagnosis: ‘DD inflammation DD metastasis’ -> FALSE; ‘likely inflammatory’ -> FALSE).
  - A hypermetabolic finding is clearly attributed to a non-malignant cause (e.g., inflammation, infection, post-surgical changes, physiological uptake).
  - There is no mention of findings suspicious for metastasis in that location.
  - The report explicitly states the lesion is unlikely to be metastasis.
  - **Do not consider any information about metastasis, which is from the case history (‘Anamnese’). E.g., it might be mentioned that the patient has a metastasis of an organ that is not part of the current imaging exam. Do NOT consider this information. Only focus on what is seen in the current imaging exam.**

**Distinguishing Primary Tumor vs. Metastasis:**

- Progression, recurrence, or residual disease of the known primary tumor at its original site is NOT metastasis for the purpose of this task.
- A hypermetabolic focus at the site of the known primary tumor should generally be considered related to the primary, UNLESS the report explicitly suggests it represents a metastatic deposit separate from the main primary mass or indicates metastatic spread within the same organ but distinct from the primary focus.
- If the radiologist describes only the primary tumor location and its characteristics (even if hypermetabolic), do NOT label this as a metastatic location.
- Keep in mind that progression of a primary tumor in this region is NOT a metastasis.
- The lesion of the primary tumor is NOT a metastasis. Pay attention to whether the radiologist labels the lesion as a metastasis or as the primary tumor.
- If the radiologist describes the location of the primary tumor only, do NOT use this as a location for metastasis, unless the patient has metastasis in the same location as the primary tumor.

##### Extraction of Response to Treatment

**1. Guiding Principles**

This guide classifies radiology reports based on cancer treatment response. The core logic depends on the dynamic described in the report’s comparison to a prior scan: is the overall tumor burden shrinking, stable, or growing?

- **RESPONSE (R):** The direct result of successful treatment. The report describes the **elimination (Complete Response)** or **significant reduction (Partial Response)** of tumor burden, OR it **confirms the continued absence of disease** after a prior complete response.
- **STABLE DISEASE (SD):** A state of equilibrium. This applies **only when a measurable tumor is present** and the report describes it as not having significantly changed in size *since the last scan*.
- **PROGRESSION (PD):** The cancer is growing. The report describes that existing tumors are bigger or, critically, that new tumors have appeared.
- **NOT APPLICABLE (NA):** The scan is not for treatment follow-up.

**If the radiologist uses ‘RESPONSE’, ‘STABLE DISEASE’, ‘PROGRESSION’ according to criteria like RECIST or RANO, always extract the radiologists conclusion directly**.

**2. Classification Criteria**

RESPONSE (R - Ansprechen / Remission)

- **Definition:** The report documents a positive effect of treatment by describing a clear reduction or elimination of tumor burden, or the continued absence of disease, *compared to the immediate previous examination*.
- **Applies When:**
  - **Complete Response:** The report states that all known tumors have disappeared.
  - **Partial Response:** The report states that known tumors have significantly decreased in size.
  - **Continued Remission:** The report confirms that a patient who previously achieved a Complete Response remains free of disease.
  - **Post-Surgical/Interventional:** The report is the first one after a major intervention (like surgery) and documents the successful removal or reduction of a tumor, even if other tumors remain.
- **Key Conditions:** No new cancerous lesions are described.
- **Look For (Keywords):** ‘komplette/partielle Remission’, ‘deutliche Größenabnahme’, ‘signifikante Regredienz’, ‘gutes Ansprechen’, ‘kein Tumornachweis mehr’, ‘Zustand nach Resektion’, ‘kein Anhalt für Rezidiv’.

STABLE DISEASE (SD - Stabile Erkrankung)

- **Definition:** The report documents that a **known, measurable tumor is not changing significantly** *compared to the immediate previous examination*.
- **Applies When**
  - The report describes the size of existing tumor(s) as unchanged since the last scan.
  - The report describes a tumor as stable compared to the last scan, even if the clinical history mentions it was much larger in the distant past (e.g., a tumor shrank from 5cm to 2cm months ago, and is now described as ‘stable at 2cm’). The stability is the most recent event.
- **Key Conditions:** No new cancerous lesions are described. The patient must have measurable disease.
- **Look For (Keywords):** ‘stabile Erkrankung’, ‘unveränderter Befund’, ‘konstante Läsionsgröße’, ‘Befundstabilität’, ‘keine wesentliche Dynamik’.

PROGRESSION (PD - Progrediente Erkrankung)

- **Definition:** The report documents an overall increase in tumor burden.
- **Applies When:**
  - The report states that existing tumors have significantly **increased in size.**
  - **The report mentions that one or more new cancerous lesions have appeared.**
- **Key Conditions:** The appearance of new lesions signifies PROGRESSION, even if other tumors are stable or shrinking.
- **Look For (Keywords):** ‘progrediente Erkrankung’, ‘Progression’, ‘Größenzunahme’, ‘Befundverschlechterung’, and especially: **‘neue Läsionen’**, **‘neu aufgetretene Metastasen’**.

NOT APPLICABLE (NA)

- **Definition:** The scan cannot be used to assess treatment response.
- **Applies When:** It is a baseline/staging scan (Erststaging), no prior scan is available for comparison (keine Voruntersuchung), or the clinical question is unrelated to cancer follow-up.

**3. Examples for Classification**

Use these self-contained examples to guide classification logic.

**Example 1**

- **SCENARIO:** A patient undergoes chemotherapy for multiple lung metastases.
- **REPORT FINDING:** “Compared to the prior study, all previously described lung metastases have resolved.”
- **CLASSIFICATION:** RESPONSE
- **REASONING:** The report describes the complete disappearance of the tumor, which is a Complete Response.

**Example 2**

- **SCENARIO:** A patient who previously had a complete response has a routine 6-month follow-up scan.
- **REPORT FINDING:** “No evidence of tumor recurrence. Post-operative changes are stable compared to the prior examination.”
- **CLASSIFICATION:** RESPONSE
- **REASONING:** The key finding is the continued absence of disease. It cannot be Stable Disease because there is no measurable tumor. This confirms a continued state of remission.

**Example 3**

- **SCENARIO:** A patient is being treated for a 5 cm liver metastasis.
- **REPORT FINDING:** “The known liver metastasis has decreased in size and now measures 2 cm in diameter. This represents a partial response to therapy.”
- **CLASSIFICATION:** RESPONSE
- **REASONING:** The report explicitly describes a significant reduction in tumor size.

**Example 4**

- **SCENARIO:** A patient who previously had a partial response (tumor shrank to 2 cm) has a follow-up scan 3 months later.
- **REPORT FINDING:** “The 2 cm liver metastasis is unchanged in size compared to the study from 3 months ago.”
- **CLASSIFICATION:** STABLE DISEASE
- **REASONING:** The most recent event described is stability. The classification is based on the finding in the most recent interval, not the entire treatment history.

**Example 5**

- **SCENARIO:** A patient with a primary tumor and a single metastasis undergoes surgery to remove the primary tumor. This is the first scan after the operation.
- **REPORT FINDING:** “Status post-resection of the primary tumor in the colon. The known liver metastasis is unchanged.”
- **CLASSIFICATION:** RESPONSE
- **REASONING:** The scan documents the result of the major therapeutic intervention (surgery), which successfully removed a large part of the tumor burden.

**Example 6**

- **SCENARIO:** A patient from the previous example has another follow-up scan 6 months later.
- **REPORT FINDING:** “The known liver metastasis remains stable in size compared to the prior post-operative scan. No new lesions are seen.”
- **CLASSIFICATION:** STABLE DISEASE
- **REASONING:** The focus is now on the dynamic of the remaining, measurable tumor. Since it is unchanged, the current state is stable.

**Example 7**

- **SCENARIO:** A patient is on active treatment for known lesions.
- **REPORT FINDING:** “The known liver lesions are stable, but a new 1 cm lesion is now visible in the spleen, suspicious for a metastasis.”
- **CLASSIFICATION:** PROGRESSION
- **REASONING:** The appearance of a new lesion is the strongest indicator of progression and overrides stability elsewhere.

**Example 8**

- **SCENARIO:** A patient is on active treatment for known lesions in the liver and spleen.
- **REPORT FINDING:** “The known liver lesions have decreased in size, but the lesion of the spleen has significantly increased in size.”
- **CLASSIFICATION:** PROGRESSION
- **REASONING:** The growth of an existing lesion overrides decrease elsewhere.

**Example 9**

- **SCENARIO:** A patient with astrocytoma had resection of the primary tumor and has a post-op scan.
- **REPORT FINDING:** “No evidence for tumor-lesions. ‘Stable Disease’ according to RANO criteria.”
- **CLASSIFICATION:** STABLE DISEASE
- **REASONING:** The key finding is the classification of the radiologist as stable disease. This overrides anything else.

**4. Final Rules & Prioritization**

1. **If the radiologist makes a clear classification, this is king and should be extracted.**
2. The appearance of a **new lesion characterized as likely malignant** is the strongest signal for PROGRESSION. If the radiologist suggests a non-malignant cause (e.g., infection, inflammation) as more likely, it should not be classified as progression based on that finding alone.
3. **No Tumor ≠ Stable Disease:** A state of “no detectable tumor” is RESPONSE, UNLESS the radiologist explicitly categorizes the finding as STABLE DISEASE.
4. **Focus on the Most Recent Change:** The classification must reflect the dynamic described in the current report (e.g., ‘stable compared to last scan,’ ‘shrinking since last scan’). The primary classification comes from the most recent interval change.

##### Extraction of radiologically tumor free

- If you have chosen COMPLETE_RESPONSE or NOT_APPLICABLE for PET-CT or RESPONSE or NOT_APPLICABLE for non-PET imaging also indicate whether the patient is completely tumor free, in other words, there is no evidence of any tumor (‘Kein Tumornachweis’).
- If you have chosen PARTIAL_RESPONSE or STABLE_DISEASE or PROGRESSION always answer with false.
- If you chose COMPLETE_RESPONSE based on metabolic finding, always choose yes.
- If the imaging exam failed for any reason, answer with yes.
- Do not consider any information about metastasis, which is from the case history (‘Anamnese’). Only focus on what is seen in the current imaging exam.

#### Prompts

These are prompts used in the classification pipelines. The prompts are better understood in the conext of the code. We encourage the reader to examine the code together with the prompts [here](https://github.com/scjohannes/llm-med-extraction).

##### Metastasis Classification

###### System Prompt

You are a highly trained medical AI assistant helping users to extract the location of metastases from radiology reports.

###### Prompts 1-19

This template contains the instructions common to all functions. Placeholders {region_name} and {metastasis_questions} are dynamically replaced based on the specific body region(s) being evaluated.:

**ROLE INSTRUCTIONS:**
“Assume you are a trained and experienced radiologist analyzing imaging scans (CT, MRI, X-Ray, or CT/PET) from cancer patients.
You will be given a report of the imaging exam of the {region_name} region(s) of a patient.
You are asked to extract whether this patient presently has metastasis mentioned in the report.
Please answer for each of the following separately:

{metastasis_questions}

**GENERAL:**
Interpreting PET-CT Findings:

- Pay close attention to descriptions of lesions, particularly those identified as 'hypermetabolic', 'FDG-avid', having 'increased uptake', or showing a specific 'SUV' (Standardized Uptake Value). These indicate metabolic activity often associated with cancer but require careful interpretation within the report's context.

- Radiologists may use terms like 'lesion', 'nodule', 'mass', 'uptake', or 'activity' instead of explicitly stating 'metastasis'. Your task is to determine if the description and context imply metastatic disease.

Interpreting CT and MRI-Findings:

- Identify if the radiologist explicitly labels any finding as 'Metastase','metastasenverdächtig', or similar.

- Assess if the radiologist describes lesions/nodules/masses with characteristics and then explicitly states they are suspicious for, concerning for, consistent with, likely, or typical of metastasis.

- Note if the radiologist offers a differential diagnosis; the likelihood assigned to metastasis is key (see TRUE/FALSE criteria).

**ANSWERING TRUE/FALSE:**
Answer with TRUE if there is:

- A clear mention of metastasis in the specific location.

- A description of a lesion where the radiologist expresses a *strong suspicion* or high likelihood of it representing metastasis (e.g., 'metastasenverdächtig', 'concerning for metastatic deposit', 'vereinbar mit Metastasierung', 'wahrscheinliche Metastasen').

Answer with FALSE if:

- Metastasis is mentioned but considered *less likely* than other possibilities (e.g., listed lower        in the differential diagnosis: 'DD inflammation DD metastasis' -> FALSE; 'likely inflammatory' -> FALSE).

- A hypermetabolic finding is clearly attributed to a non-malignant cause (e.g., inflammation, infection, post-surgical changes, physiological uptake).

- There is no mention of findings suspicious for metastasis in that location.

- The report explicitly states the lesion is *unlikely* to be metastasis

- Do not consider any information about metastasis, which is from the case history ('Anamnese'). E.g., it might be mentioned that the patient has a metastasis of an organ that is not part of the current imaging exam. Do NOT consider this information. Only focus on what is seen in the current imaging exam.

**DISTINGUISHING PRIMARY TUMOR VS. METASTASIS:**

- Progression, recurrence, or residual disease of the known primary tumor at its original site is NOT metastasis for the purpose of this task.

- A hypermetabolic focus at the site of the known primary tumor should generally be considered related to the primary, UNLESS the report explicitly suggests it represents a metastatic deposit separate from the main primary mass or indicates metastatic spread within the same organ but distinct from the primary focus.

- If the radiologist describes only the primary tumor location and its characteristics (even if hypermetabolic), do NOT label this as a metastatic location.

- Keep in mind that progression of a primary tumor in this region is NOT a metastasis.

- The lesion of the primary tumor is NOT a metastasis. Pay attention to whether the radiologist labels the lesion as a metastasis or as the primary tumor.

- If the radiologist describes the location of the primary tumor only, do NOT use this as a location for metastasis, unless the patient has metastasis in the same location as the primary tumor:

The imaging report text is below this line:

-------------------------------------------

{text_placeholder}

P1-19. Depending on {region_name}, only some of {metastasis_questions} will be asked. Each is answered true or false.

.Q_CNS <- "This patient has metastasis of the brain parenchyma."
.Q_MENINGEAL <- "This patient has meningeal metastasis."
.Q_BONE <- "This patient has bone metastasis."
.Q_LYMPH <- "This patient has lymph node metastasis."
.Q_SOFT_TISSUE <- "This patient has soft tissue metastasis."
.Q_LIVER <- "This patient has liver metastasis."
.Q_ADRENAL <- "This patient has adrenal metastasis."
.Q_KIDNEY <- "This patient has kidney metastasis."
.Q_SPLEEN <- "This patient has spleen metastasis."
.Q_PANCREAS <- "This patient has pancreatic metastasis."
.Q_PERITONEAL <- "This patient has peritoneal metastasis."
.Q_OVARIAN <- "This patient has ovarian metastasis."
.Q_LUNG <- "This patient has lung metastasis."
.Q_PLEURA <- "This patient has pleural metastasis."
.Q_OTHER_ORGAN <- "This patient has metastasis in any organ mentioned."
.Q_JUSTIFICATION <- "Provide justification based only on the text."

Output example JSON only for thorax region:

{

"lung_metastasis": true,

"pleural_metastasis": false,

"lymph_node_metastasis": true,

"bone_metastasis": false,

"soft_tissue_metastasis": false,

"justification": "Multiple pulmonary nodules ‘suspicious for metastases’ and enlarged mediastinal nodes ‘concerning for metastatic disease’; no pleural deposits described."

}

##### Response to Treatment Classification

System Prompt

You are a highly trained medical AI assistant helping users to extract information on treatment response from radiology reports.

(Step 1) Prompt assessing whether report is in-scope.

Analyze the provided German imaging report text and determine if its primary purpose is to assess response to a treatment of the cancer (i.e., a follow-up after treatment, surveillance after remission, or response assessment scan).
INPUT: Full text of a German imaging report.
OUTPUT: Classify the report's purpose into ONE of the following two categories:
1. `IN_SCOPE`: The report is a follow-up, monitoring, or treatment response assessment scan. If the patient was treated in the past and we are interested in remission or relapse, this is also in scope.
2. `OUT_OF_SCOPE`: The report is for an initial diagnosis or initial staging (`Baselinestaging`), the exam didn't take place, the scan was done to rule out infection or anything else unrelated, or the scan was clearly done before any anti-cancer treatment including surgery was begun.
Below this line starts the radiology report:
---------------------------------------

If RANO or RECIST classification was used in the report (Step 2 == YES).

You are given a german radiology report for a cancer patient. Response to treatment was classified using RANO or RECIST. Extract ad verbatim the sentence in which the radiologist does the OVERALL reponse classification (Gesamtauswertung) according to RANO or RECIST.
Below this line starts the radiology report:
---------------------------------------

If the report is from a PET scan (Step 3 == YES).

```
Now map verbatim response to one PET-CT metabolic response category.
Must be one of: ['COMPLETE_RESPONSE','PARTIAL_RESPONSE','STABLE_DISEASE','PROGRESSION']
MAPPING HINTS:
- 'Complete Response', 'Komplette Remission' -> COMPLETE_RESPONSE
- 'Partial Response', 'Partielle Remission' -> PARTIAL_RESPONSE
- 'Stable Disease', 'Stabile Erkrankung'     -> STABLE_DISEASE
- 'Progression', 'Progrediente Erkrankung'   -> PROGRESSION
Additionally, determine if the patient is completely tumor free.
- Set `no_tumor` to `TRUE` ONLY IF your classification is `COMPLETE_RESPONSE` AND the full report text (from the previous turn) indicates the patient is completely tumor free ('Kein Tumornachweis').
- Set `no_tumor` to `FALSE` in all other cases.
```

If the report is not from a PET scan (Step 3 == No).

```
Now map the verbatim response to one OVERALL response category.
Must be one of: ['RESPONSE','STABLE_DISEASE','PROGRESSION'].
MAPPING HINTS:
- 'Complete Response', 'Komplette Remission' -> RESPONSE
- 'Partial Response', 'Partielle Remission' -> RESPONSE
- 'Stable Disease', 'Stabile Erkrankung'     -> STABLE_DISEASE
- 'Progression', 'Progrediente Erkrankung'   -> PROGRESSION
Additionally, determine if the patient is completely tumor free.
- Set `no_tumor` to `TRUE` ONLY IF your classification is `RESPONSE` AND the full report text (from the previous turn) indicates the patient is completely tumor free ('Kein Tumornachweis').
- Set `no_tumor` to `FALSE` in all other cases.
```

If RANO or RECIST classification was not used in the report (Step 2 == NO) and the report is a PET scan (Step 3 == YES).

First task: Analyze & extract text without structured reponse. Second task: Map to pre-specified classification. Third task: Answering tumor free? TRUE/FALSE

<first task>
Analyze the provided German PET-CT report text and classify the patient's cancer status based strictly on described changes in metabolic activity (FDG uptake).Ignore changes in size (CT findings) unless metabolic information is entirely absent.

INPUT: Full text of a German PET-CT report.
OUTPUT: Classify the report into ONE of the following categories:
1. `COMPLETE_RESPONSE`
2. `PARTIAL_RESPONSE`
3. `STABLE_DISEASE`
4. `PROGRESSION`
CLASSIFICATION CRITERIA (Focus ONLY on Metabolic Activity):
1. `COMPLETE_RESPONSE` (CR - Komplette Metabolische Remission):
General: Report indicates *complete resolution* of abnormal metabolic activity in all known lesions, down to background levels. No FDG-avid malignant lesions remain.
Solid Tumors: Look for terms explicitly stating complete metabolic response (e.g., 'komplette metabolische Remission,' 'kein pathologischer Hypermetabolismus mehr nachweisbar,' 'vollständige Normalisierung des Stoffwechsels'). This also includes COMPLETE surgical resection of the tumor and, if existing, its metastases.
Lymphoma/Blood Cancer: Look for'Deauville Score 1,' 'Deauville Score 2,' or 'Deauville Score 3' as the final assessment for all originally involved sites, AND absence of new FDG-avid lesions. Terms like 'komplette metabolische Remission.'
Note: Residual non-avid findings on CT do not preclude CR.
2. `PARTIAL_RESPONSE` (PR - Partielle Metabolische Remission):
General: Report indicates a significant decrease in metabolic activity (FDG uptake/SUV) in known lesions compared to the prior scan, but some residual abnormal uptake persists (above background levels).
Solid Tumors: Look for terms indicating partial metabolic response (e.g., 'partielle metabolische Remission,' 'deutliche Abnahme/Reduktion der Stoffwechselaktivität/des SUV,' 'deutliche Regredienz des Hypermetabolismus,' but not complete resolution). This also includes INcomplete surgical resection of the tumor or its metastases.
Lymphoma/Blood Cancer: May be indicated by a shift to a lower Deauville Score (e.g., from 5 to 4), or descriptions like 'deutliche Abnahme der metabolischen Aktivität' but uptake still clearly above background/liver (i.e., still DS 4 or 5 but improved).Use this category if response is clear but doesn't meet CMR criteria (DS 1-3).
Note: Significant metabolic decrease defines PMR, even if size is stable or slightly increased.
3. `STABLE_DISEASE` (SD - Stabile Metabolische Erkrankung):
General: Report indicates no significant change (neither significant decrease nor increase) in metabolic activity (FDG uptake/SUV values) in known lesions compared to the prior scan.
Solid Tumors: Look for terms like 'stabile metabolische Aktivität,' 'unveränderter SUV,' 'keine signifikante Änderung des Hypermetabolismus,' 'stabiler Befund.'
Lymphoma/Blood Cancer:Often indicated by a persistent 'Deauville Score 4' or 'Deauville Score 5' without mention of significant change compared to prior or new lesions. Look for terms like 'persistierende metabolische Aktivität', 'stabile Erkrankung.'
4. `PROGRESSION` (PD - Progrediente Metabolische Erkrankung):
General: Report indicates a significant *increase* in metabolic activity (FDG uptake/SUV) in existing lesions OR, critically, the appearance of new FDG-avid lesions consistent with malignancy.
Solid Tumors: Look for terms indicating metabolic progression (e.g., 'metabolische Progression,' 'Zunahme der Stoffwechselaktivität/des SUV,' 'progredienter Hypermetabolismus'). Crucially, look for 'neue Herde,' 'neue Läsionen,' 'neue stoffwechselaktive Läsionen.'
Lymphoma/Blood Cancer: Look for 'Deauville Score 4' or 'Deauville Score 5' with context indicating worsening compared to prior scan, OR any mention of 'neue FDG-avide Läsionen,' 'neue Herde'(often implicitly DS 5). Also terms like 'Progression,' 'progrediente Erkrankung. '
Note: New metabolically active lesions automatically mean `PROGRESSION`. Increased metabolic activity in existing lesions also means `PROGRESSION`, even if size decreases.
PRIORITIZATION:
-	Metabolic findings (PET) override anatomical findings (CT).
-	The 'Beurteilung' (Impression/Conclusion) section often contains the final summary but verify against the 'Befund' (Findings) and 'Vergleich' (Comparison) sections.
-	The presence of new, metabolically active lesions consistent with malignancy is the strongest indicator for `PROGRESSION`.
-	Explicit mention of Deauville Scores (for lymphoma) is a primary classification driver. Distinguish carefully between DS 1-3 (CMR), and DS 4/5 (can be PMR, SMD, or PMD depending on comparison/new lesions).
Below this line starts the radiology report:
 ---------------------------------------
</first task>

<second task>
Now please map your final decision to the following STRUCTURE and return ONLY the structured object:
- response_to_trt_pet: one of['COMPLETE_RESPONSE','PARTIAL_RESPONSE','STABLE_DISEASE','PROGRESSION']
- justification: short 1-2 sentence rationale.
</second task>
<third task>
If you have chosen `COMPLETE_RESPONSE` you are also asked to indicate whether the patient is completely tumor free, in other words, there is no evidence of any tumor ('Kein Tumornachweis').
If you have chosen `PARTIAL_RESPONSE` or `STABLE_DISEASE` or `PROGRESSION` always answer with false.
OUTPUT: no_tumor: TRUE / FALSE
</third task>

If RANO or RECIST classification was not used in the report (Step 2 == NO) and the report is not a PET scan (Step 3 == No).

```
<first task>
Analyze the provided German CT or MRI report text and classify the patient's cancer status.
INPUT: Full text of a German CT or MRI report.
OUTPUT: Classify the report into ONE of the following categories:
1.  `RESPONSE`
2.  `STABLE_DISEASE`
3.  `PROGRESSION`
GUIDING PRINCIPLES
The core logic depends on the dynamic described in the report's comparison to a prior scan: is the overall tumor burden shrinking, stable, or growing?
RESPONSE (R): The direct result of successful treatment. The report describes the elimination (Complete Response) or significant reduction (Partial Response) of tumor burden, OR it confirms the continued absence of disease after a prior complete response.
STABLE DISEASE (SD): A state of equilibrium. This applies only when a measurable tumor is present and the report describes it as not having significantly changed in size since the last scan.
PROGRESSION (PD): The cancer is growing. The report describes that existing tumors are bigger or, critically, that new tumors have appeared.
CLASSIFICATION CRITERIA
1.`RESPONSE` (R - Ansprechen / Remission):
General: Report indicates either complete disappearance of all known/target lesions OR a significant decrease in the size (e.g., diameter, volume) of known/target lesions compared to the prior scan. No new lesions are present, and no unequivocal progression of non-target lesions. This category covers both complete and substantial partial responses. A scan performed to confirm the outcome of a recent surgery that successfully removed all visible disease is also classified as RESPONSE for that event.
Look For: Terms explicitly stating complete or partial response, or significant size reduction/disappearance (e.g., 'komplette Remission,' 'partielle Remission,' 'deutliche Größenabnahme der Läsionen,' 'signifikante Regredienz,' 'vollständige Rückbildung aller Läsionen,' 'kein Tumornachweis mehr,' 'Läsionen nicht mehr abgrenzbar/nachweisbar,' 'vollständige narbige Ausheilung,' 'größenregredient,' 'deutliche Verkleinerung,' 'gutes Ansprechen'). Comparison to a previous scan must confirm disappearance or significant reduction.
2. `STABLE_DISEASE` (SD - Stabile Erkrankung):
General: Report indicates no significant change (neither sufficient reduction for `RESPONSE` nor sufficient increase/new lesions for `PROGRESSION`) in the size or characteristics of known lesions compared to the prior scan.
Look For: Terms like 'stabile Erkrankung,' 'unveränderter Befund,' 'konstante Läsionsgröße,' 'keine signifikante Größenänderung,' 'Befundstabilität,' 'keine wesentliche Dynamik.'
3.  `PROGRESSION` (PD - Progrediente Erkrankung / Progression):
General: Report indicates a significant increase in the size of existing lesions, OR, critically, the appearance of new lesions consistent with malignancy, OR unequivocal progression of non-target lesions.
Look For: Terms indicating progression (e.g., 'progrediente Erkrankung,' 'Progression,' 'Größenzunahme der Läsionen,' 'Befundverschlechterung,' 'größenprogredient').
Crucially, look for any mention of 'neue Läsionen,' 'neue Herde,' 'neu aufgetretene Metastasen/Filiae,' 'neu abgrenzbare Läsionen.' The appearance of new lesions typically signifies progression regardless of the behavior of previously known lesions. The identification of a recurrence (e.g., 'Rezidiv', 'Tumorrezidiv', 'Lokalrezidiv') after a period of remission or after surgical resection is a definitive sign of progression.
EXAMPLES FOR CLASSIFICATION
Use these self-contained examples to guide classification logic.
Example 1
SCENARIO: A patient undergoes chemotherapy for multiple lung metastases.
REPORT FINDING: 'Compared to the prior study, all previously described lung metastases have resolved.'
CLASSIFICATION: RESPONSE
REASONING: The report describes the complete disappearance of the tumor, which is a Complete Response.
Example 2
SCENARIO: A patient who previously had a complete response has a routine 6-month follow-up scan.
REPORT FINDING: 'No evidence of tumor recurrence. Post-operative changes are stable compared to the prior examination.'
CLASSIFICATION: RESPONSE
REASONING: The key finding is the continued absence of disease. It cannot be Stable Disease because there is no measurable tumor. This confirms a continued state of remission.
Example 3
SCENARIO: A patient is being treated for a 5 cm liver metastasis.
REPORT FINDING: 'The known liver metastasis has decreased in size and now measures 2 cm in diameter. This represents a partial response to therapy.'
CLASSIFICATION: RESPONSE
REASONING: The report explicitly describes a significant reduction in tumor size.
Example 4
SCENARIO: A patient who previously had a partial response (tumor shrank to 2 cm) has a follow-up scan 3 months later.
REPORT FINDING: 'The 2 cm liver metastasis is unchanged in size compared to the study from 3 months ago.'
CLASSIFICATION: STABLE DISEASE
REASONING: The most recent event described is stability. The classification is based on the finding in the most recent interval, not the entire treatment history.
Example 5
SCENARIO: A patient with a primary tumor and a single metastasis undergoes surgery to remove the primary tumor. This is the first scan after the operation.
REPORT FINDING: 'Status post-resection of the primary tumor in the colon. The known liver metastasis is unchanged.'
CLASSIFICATION: RESPONSE
REASONING: The scan documents the result of the major therapeutic intervention (surgery), which successfully removed a large part of the tumor burden.
Example 6
SCENARIO: A patient from the previous example has another follow-up scan 6 months later.
REPORT FINDING: 'The known liver metastasis remains stable in size compared to the prior post-operative scan. No new lesions are seen.'
CLASSIFICATION: STABLE DISEASE
REASONING: The focus is now on the dynamic of the remaining, measurable tumor. Since it is unchanged, the current state is stable.
Example 7
SCENARIO: A patient is on active treatment for known lesions.
REPORT FINDING: 'The known liver lesions are stable, but a new 1 cm lesion is now visible in the spleen, suspicious for a metastasis.'
CLASSIFICATION: PROGRESSION
REASONING: The appearance of a new lesion is the strongest indicator of progression and overrides stability elsewhere.
Example 8
SCENARIO: A patient is on active treatment for known lesions in the liver and spleen.
REPORT FINDING: 'The known liver lesions have decreased in size, but the lesion of the spleen has significantly increased in size.'
CLASSIFICATION: PROGRESSION
REASONING: The growth of an existing lesion overrides decreases elsewhere.
Example 9
SCENARIO: A patient with astrocytoma had resection of the primary tumor and has a post-op scan.
REPORT FINDING: 'No evidence for tumor-lesions.'
CLASSIFICATION: STABLE DISEASE
REASONING: The key finding is the classification of the radiologist as stable disease. This overrides anything else.
FINAL RULES
-   Prioritize Overriding Information: Pay close attention to the entire report, including addenda, corrections, or later findings (e.g., 'Zusatzbefund', 'Nachtrag', 'Korrigendum'). Information in these sections may provide critical context that overrides or re-interprets the main assessment. The final classification must be based on the most complete and final context provided.
-   The appearance of a new lesion characterized as likely malignant is a strong signal for PROGRESSION.
-   If the radiologist suggests a non-malignant cause (e.g., infection, inflammation) as more likely, it should not be classified as progression based on that finding alone.
-   No Tumor ≠ Stable Disease: A state of 'no detectable tumor' is RESPONSE, UNLESS the radiologist explicitly categorizes the finding as STABLE DISEASE.
-   Focus on the Most Recent Change: The classification must reflect the dynamic described in the current report (e.g., 'stable compared to last scan,' 'shrinking since last scan'). The primary classification comes from the most recent interval change.
Below this line starts the radiology report:
  ---------------------------------------
</first task>
```

Prompt for structured output

<second task>
Now please map your final decision to the following STRUCTURE and return ONLY the structured object - extract_treatment_response_non_pet: one of ['RESPONSE','STABLE_DISEASE','PROGRESSION']
- justification: short 1-2 sentence rationale.
</second task>
<third task>
If you have chosen `RESPONSE` you are also asked to indicate whether the patient is completely tumor free, in other words, there is no evidence of any tumor ('Kein Tumornachweis').
If you have chosen `STABLE_DISEASE` or `PROGRESSION` always answer with false.
OUTPUT: no_tumor: TRUE / FALSE
</third task>

##### Diagnosis Classification

System prompt

"You are a highly trained medical AI assistant helping users to match free text diagnoses to a list of diagnosis categories provided by the user."

Free text extraction prompt

Please extract the first oncology diagnosis from the <Anamnese> section of the following german radiology report ad verbatim. Don't extract anything else.
The imaging report text is below this line:
-------------------------------------------
{text_placeholder}

Prompt for structured output

Your task is to read the provided medical text, which is a free text diagnosis from an Electronic Health Record (EHR).
The input text will most likely be in German. Identify the primary diagnosis from the text.

Assign the free text diagnosis to the best matching diagnosis category from the following list:

"Primary Brain Tumor",
"Head and Neck Tumors",
"Thyroid Cancer",
"Lung Tumor (including Mesothelioma)",
"Breast Cancer",
"Gastric Cancer",
"Esophageal Cancer",
"Pancreatic Cancer",
"Cholangiocarcinoma",
"Hepatocellular Carcinoma",
"Small Bowel Cancer",
"Colorectal Cancer",
"Kidney Tumor",
"Cancer of the Urinary Tract (including Ureter and Bladder)",
"Prostate Cancer",
"Gynecological Tumors",
"Primary Skin Tumor (including Melanoma and Non-Melanoma)",
"Sarcoma",
"Lymphoma",
"Multiple Myeloma",
"Leukemia",
"Neuroendocrine Tumors",
"Testicular Cancer",
"Cancer of Unkown Primary",
"Other",
"Unclear",
"No Malignant Disease",
"Not Applicable"

Choose 'Unclear' if the diagnosis is uncertain or not clearly stated.
Choose 'No Malignant Disease' if the text indicates no malignancy.
Prioritize the most specific diagnosis mentioned in the text.
If the diagnosis is not clear and multiple differential diagnoses are listed (often abbreviated with 'DD') choose the first differential diagnosis listed.

Always provide the best matching category from the list only and do not provide explanations.

The diagnosis text is below:
{text_placeholder}
